# Community Informed Approach for Designing Seasonal Malaria Chemoprevention Delivery in Semi-Nomadic Population in Turkana Central Sub-County, Kenya: Results from Baseline Mixed-Methods Study

**DOI:** 10.1101/2024.12.16.24319133

**Authors:** Natalya Kostandova, Catherine Kafu, Tabitha J. Chepkwony, George Ambani, Lucy Abel, Beatrice Rotich, Joseph Kipkoech Kirui, Emmah Kimachas, Emily Robie, Kaman Lowoi, David Ekai, Gilchrist Lokoel, Omalo Orinda, Suzanne Van Hulle, Robert Mgeni, Wendy Prudhomme O’Meara, Amy Wesolowski, Becky Genberg, Diana Menya

## Abstract

Following the expansion of the World Health Organization’s guidelines on eligibility for the seasonal malaria chemoprevention (SMC), the government of Kenya prepared to implement SMC for a first time in Turkana Central Sub-County in 2024. To inform design of SMC, we conducted a baseline qualitative and quantitative study. Using a stratified cluster sampling approach, we enrolled 198 households with children 6 months - 5 years old, and tested all individuals one year and older using a Rapid Diagnostic Test (RDT) and polymerase chain reaction (PCR). We carried out 82 key informant interviews, 31 focus group discussions with community health workers, elders, and caretakers of children, and 60 In-depth interviews with caretakers. Malaria prevalence was 21% [9%-42%] as reported by RDT and 21% [11%-38%] by PCR. Prevalence varied across villages, with the highest positivity of 73% by PCR. While SMC was perceived positively, challenges identified included physical access (households far from village centers, pastoralists, children living in the streets); stigma (children living with disabilities, households with members struggling with alcohol use); and acceptability (traditionalists, highly educated households). This study found that while malaria burden is high in the region, SMC may be a feasible approach to reduce its burden and transmission. However, implementation of SMC should be tailored, with a combination of centralized distribution supplemented by door-to-door and outreach to pastoralists and people in the interior. Using existing grassroots structures, such as Tree of Men and religious and group leaders, and intensive mobilization will be critical for success of this intervention.

## Introduction

Malaria is a mosquito-borne disease that can lead to serious complications including death. Despite continuous investment in malaria programs, in 2021, there were an estimated 247 million cases and 619 thousand deaths globally, with 95% of cases and 96% of deaths occurring in the World Health Organization (WHO) African Region [1]. Most malaria control strategies have relied on a combination of vector control using insecticide-treated nets (ITNs) and indoor residual spraying (IRS), and case management to correctly diagnose and treat infections [2,3]. In addition, multiple prevention strategies for particular high-risk groups, such as intermittent preventive treatment of malaria in pregnancy (IPTp) and seasonal malaria chemoprevention (SMC), are deployed [3].

Until 2022, SMC was administered according to WHO guidelines released in 2012. Under these older guidelines, the WHO has recommended the use of SMC, which consisted of monthly administration of antimalaria medicines during the season of greatest malaria risk [4], as a part of malaria control in highly seasonal transmission areas of the Sahel sub-region in Africa [2]. In most settings, SMC involved the administration of a full treatment course of anti-malaria drugs, most often a combination of sulfadoxine-pyrimethamine plus amodiaquine (SP+AQ), administered at 4-week intervals to all children 3 – 59 months old [2]. This regimen has consistently been shown to be safe and effective [5,6]. Many countries implemented four monthly cycles, although additional cycles may be recommended depending on the transmission intensity and duration [2]. How SMC is implemented varies across settings. In some contexts, SMC is administered at a central location such as a health facility or school (called ‘fixed-points’), or door-to-door delivery, wherein health personnel, community health workers, or trained staff bring the drug to the households with eligible population [7]. Research has demonstrated that the optimal approach for drug distribution may vary from context to context [7–9].

In 2022, the WHO updated its recommendations for SMC to include all children belonging to age groups at high risk of severe malaria in areas of seasonal malaria transmission, thereby not restriction SMC to the Sahel Region, nor restricting the age range and number of recommended monthly cycles to expand who may benefit from these programs [4,8]. Due to this geographic expansion, Turkana County, a semi-arid region of Northern Kenya bordering Ethiopia and Uganda, became eligible for SMC. Historically, the environment and climate in Turkana were thought to be unsuitable for malaria transmission, and as a result, there have been limited malaria control measures deployed in this area [9]. However, recent studies demonstrated a high malaria prevalence in the region, with a large proportion of asymptomatic infections and local transmission [10,11]. In 2022, there were over 274,000 confirmed cases of malaria reported in Turkana County, with 61% of cases occurring during four months (July – October) [12].

Despite the high burden of malaria in Turkana County, the use of insecticide-treated nets and other measures of malaria control is low [10,13,14], with much of the population sleeping outdoors [14]. Turkana is a primarily rural county whose population historically relied on nomadic pastoralism as a main source of livelihood. Although recent years have seen a shift away from pastoralism due to drought, environmental degradation, and population growth, the majority of the county still depends on livestock [15]. These nomadic and semi-nomadic populations in Turkana historically have lower access to health resources, with high rates of acute malnutrition and low routine vaccination coverage [13]. Community based health campaigns, for example for measles and polio vaccination, have supplemented routine activities [16,17], sometimes targeting special populations that may be missed otherwise [18].

In 2023, The Turkana County Health Team, in collaboration with Duke and Moi Universities, and Catholic Relief Services, was preparing to implement SMC in Turkana Central Sub-county in 2024, with the vision that findings about SMC effectiveness from this phase of implementation would inform the eventual roll-out of SMC in the entirety of Turkana County [19]. Historically, large-scale distribution campaigns, such as mass drug administration for neglected tropical diseases, have documented barriers to access and specialized strategies to reach nomadic and mobile populations [20–23]. To plan and ensure that SMC programs account for population-specific challenges in Turkana Central, we carried out a mixed-methods formative study. We present the baseline, pre-SMC prevalence of malaria in the Sub-county, as well as findings from qualitative research describing the potential challenges and opportunities of SMC. Finally, we suggest strategies and adaptations to ensure high coverage and acceptability given the context in Turkana Sub-County.

## Materials and Methods

### Study setting

This was a mixed methods study conducted in all five wards of Turkana Central Sub-County of Turkana County, Kenya (Township, Kanamkemer, Kangatossa, Kerio, and Kalokol). Two communities per ward were selected using probability proportional to population size (Figure 1).

**Figure 1.**
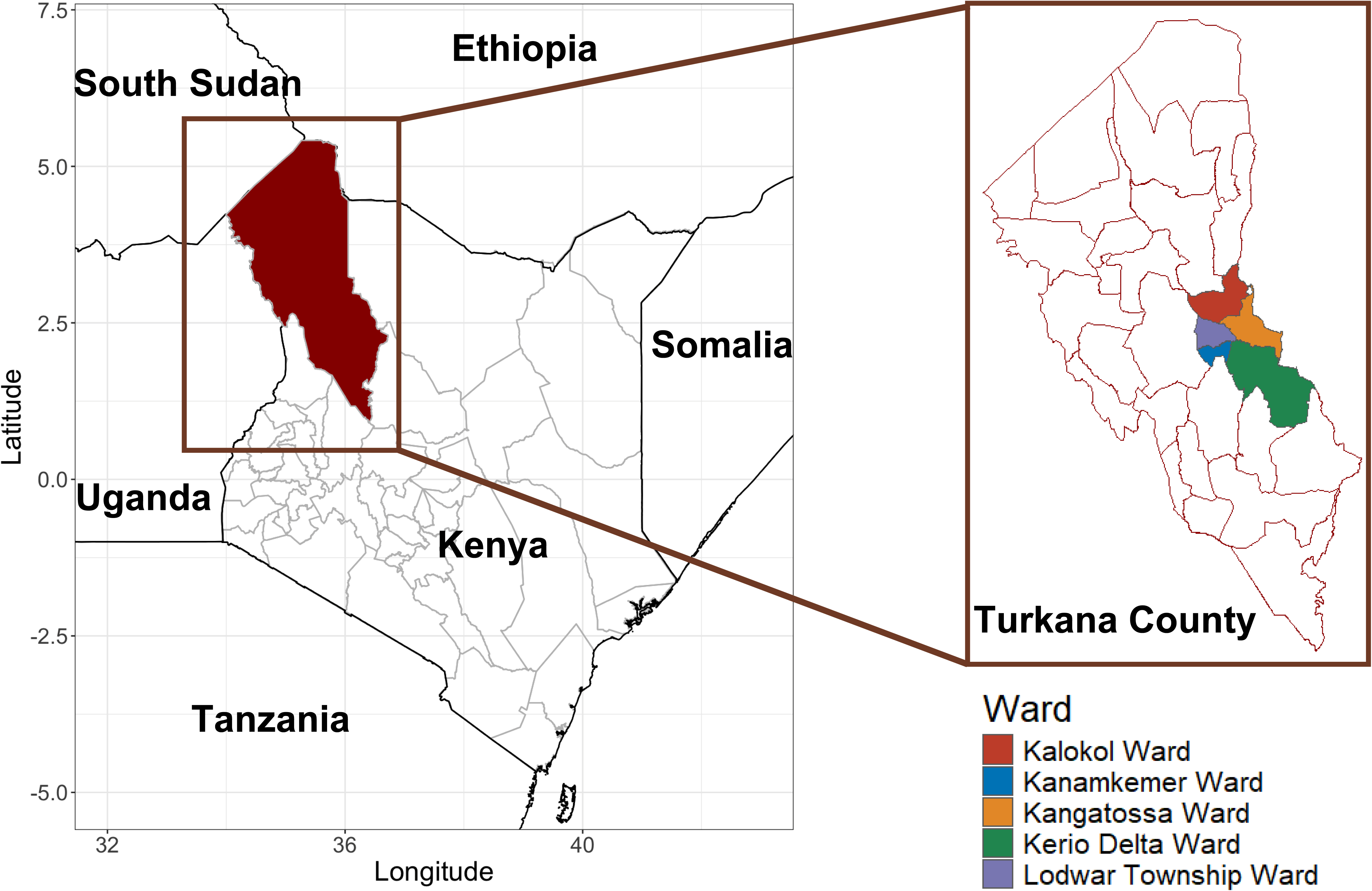
Study sites for qualitative baseline Seasonal Malaria Chemoprophylaxis study, Turkana Central Sub-County, Kenya. The map presents outlines of wards in Turkana County, with colored wards in Turkana Central Sub-County. The study took place in ten communities in this Sub-County, two per ward.

### Community entry

Prior to data collection, the research team met with representatives from government (the Ministry of Health and Sanitation Services) and non-governmental organizations (NGOs) in Lodwar to identify partners with interest and influence in the implementation of SMC in Turkana County. During these community visits, the team presented the study’s objectives to community leaders and health personnel, addressed questions about the study, and planned for the logistics of data collection. In addition, the study team aimed to understand basic characteristics of the community to inform recruitment of qualitative participants, including primary sources of livelihood, religion, and decision-making structures.

### Participant recruitment

For <u>quantitative data collection</u>, within each selected village, a sampling frame of all households with children between 6 months and 5 years old were compiled using household registers (MoH 513) which are filled, kept, and updated by Community Heath Promotors (CHPs, formerly Community Health Workers or Volunteers). In each village, up to 20 households were randomly selected that had children under 5 years old. In selected households, a household head or their proxy enumerated household members, provided information on household demographics, economic activities, recent illness and malaria prevention measures.

All household members aged 1 year and above were eligible to participate in additional survey questions, malaria rapid diagnostic testing (RDT), and dried blood spot (DBS) collection. Individuals 18 years of age or older provided written informed consent. The survey collected data on respondent demographics, practices related to medical history, travel history, and knowledge and attitudes towards malaria and malaria prevention practices (questionnaire provided in Supplementary materials Appendix 1). Children under 18 years old participated after written informed consent from a parent or guardian. Verbal assent was additionally sought from children 12 years and above.

For each household member, a malaria RDT was done and a DBS collected. When a household was not available or more than 20% of household members in a village were not contacted, a revisit was conducted. If the household was not available on the second visit, a replacement household was located. Dried blood spots were transported to Webuye, Kenya and tested for the presence of *P. falciparum* and other plasmodium species. Households with malaria positive cases by RDT were given a referral form and provided with transport money to go to the nearest health facility with their RDT results to obtain free treatment.

For <u>qualitative data collection</u>, participants aged 18 years or older were selected using a combination of purposive and snowball approaches, using different criteria for each thematic area (Table S1). Recruitment occurred iteratively such that Key Informant Interviews (KIIs) and Focus Group Discussions (FGDs) may have identified important groups for inclusion in additional data collection.

*KIIs*: In each of the ten communities, at least six community leaders were identified for KIIs on social and resource mapping. These leaders were chiefs, village, and ward administrators, religious leaders, youth leaders, village elders, and leaders of community groups like fisherfolk. To learn about the community’s health profile and challenges in the provision of health services, we interviewed key informants, including heads of health facilities, Community Health Assistants, community health mobilizers, and community health committee members.

*In-Depth Interviews (IDIs)*: Caretakers of children under five were selected to capture different experiences of seeking healthcare and access to community resources. For example, we enrolled caretakers of children living with disabilities, orphaned children, children living far away from the health facility, children who defaulted from nutrition programs or delayed vaccination, and those within families with unhealthy alcohol use.

*FGDs:* Ranking exercises to understand community member preferences for the mode of SMC administration were done separately with caregivers of children under five and with community elders. FGDs to understand travel, exploring what influenced community members’ migration, who they migrated with, and how access to healthcare changed, were carried out with members of the Tree of Men, a group of mature men that act as a decision-making mechanism for pastoralist communities, or with elders in the community. FGDs on CHP experiences were done with CHPs.

Summary of these thematic areas and profile of participants are presented in Table S1.

### Data collection

For <u>quantitative data collection</u>, interviews were carried out using KoboCollect software in Turkana or Kiswahili. Participants were enrolled from 198 households Turkana Central.

For <u>qualitative data collection</u>, semi-structured guides were created for all KIIs, IDIs, and FGDs, and were translated and back-translated from English to Kiswahili and Ngaturkana. Data collection was carried out in the language choice of participants. The exception was the mass activity domain data that were collected from participants for whom English is the primary work language; as a result, the guides were only available in English, and their interviews were carried out in English.

Discussions and interviews were recorded, unless participants did not consent to an audio recording. In these cases, only notes were taken.

In total, 410 participants were enrolled across the ten villages and Lodwar and participated in 78 KIIs, 31 FGDs and 60 IDIs.

Quantitative and qualitative data collection was carried out from August to September 2023.

### Data analysis

#### Quantitative data

Descriptive statistics for indicators of interest were calculated using *survey* package [24] in R version 4.4.1 [25] and RStudio 2024.04.2 [26], accounting for study design. At individual level, we specified stratified selection at Ward level, with three-level sampling (at Village, Household, and Household member level). Probabilities of selection were calculated at each of the three levels. For household-level variables, only two levels of selection were specified (Village and Household levels).

#### Qualitative data

Debriefings with the qualitative field research team were carried out daily. During the first two weeks of data collection, a study team member read notes from the interviews and FGDs to identify emerging key themes and criteria for the selection of participants in the upcoming days. Audio recordings were transcribed and translated directly into English verbatim. Transcripts for KIIs with community leaders and health workers were uploaded and manually coded inductively by NK using ATLAS.ti (v.23.4.0) [27] and NVivo 10 [28]. We used memos to note summary descriptions of findings from all other transcripts and notes, highlight key quotations and synthesize the results. Barriers and anticipated challenges to implementation of SMC were organized around determinants from The Designing for Behavior Change Framework. The framework is based on behavior change theories (i.e., Health Behavior Model and Theory of Reasoned Action) and is used to formalize understanding of determinants, key factors, challenges, and activities for successful design of an intervention for behavior change [29]. In our case, the desired behavior would be the uptake of SMC. The data were organized in a matrix according to the determinants of the Designing for Behavior Change Framework (i.e., perceived self-efficacy, perceived social norms, etc.), along with potential strategies to address the determinant in the implementation of the intervention. Two domains were then identified from the data that represented specific challenges to the implementation, delivery and access of SMC that cut across the determinants. Finally specific recommendations for intervention implementation informed by the context were identified.

*Ethical approval:* Study was approved by Moi University Research Ethics Committee (Approval number 0004733) and Duke University Institutional Review Board (Protocol ID Pro00115824).

## Results

### Results of the quantitative study

#### Profile of participants in quantitative study

Twenty households were enrolled in each of the ten villages in Turkana Central Sub-county, with exception of Nasingila village, where 18 households were enrolled. A total of 983 individuals were enrolled, with enrollment ranging from 72% in Atangi to 90% in Apetet (Table S2). Percentage of household members who could be found at home and were interviewed was 79%.

Median age of individuals enrolled was 12 years old [IQR: 5, 26] (Table 1). Majority of enrollees were female, and most of individuals enrolled were children of the heads of household (Table 1).

**Table 1.**
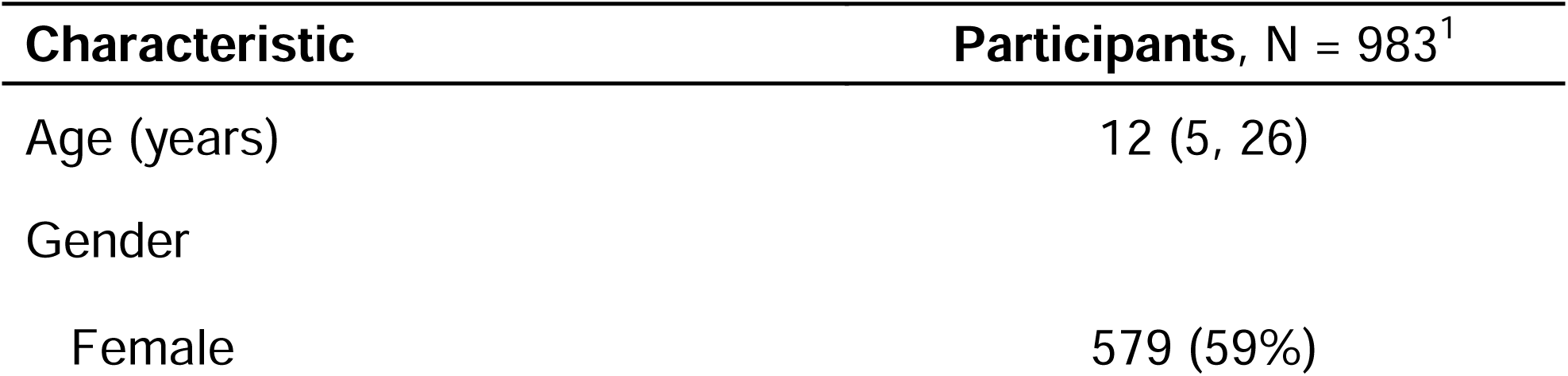
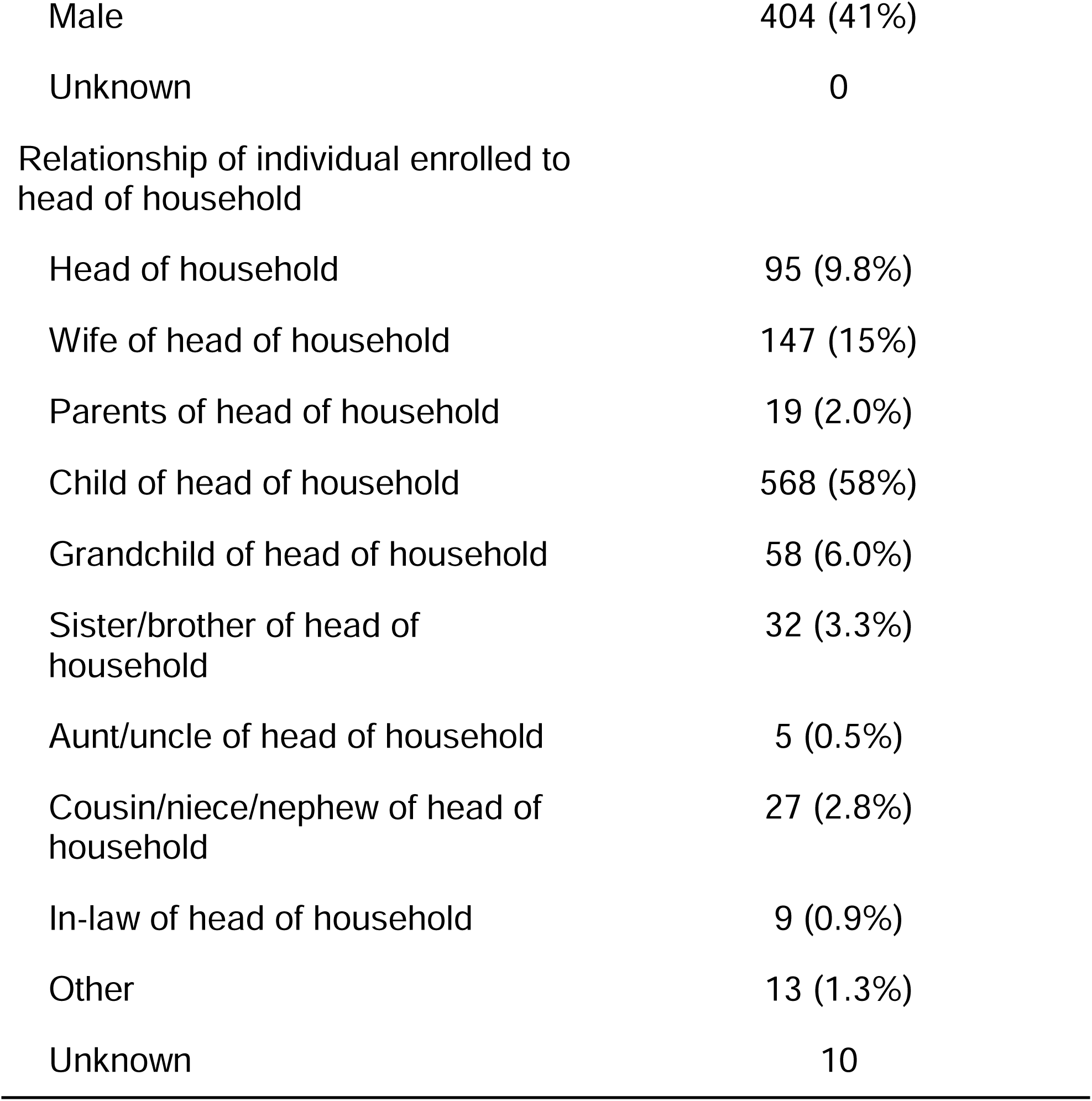
Demographic characteristics of individuals enrolled in the quantitative baseline study, Turkana Central Sub-County, 2023

At household level, there was large variability in characteristics such as primary water source and primary source of income. Wards bordering Lake Turkana reported larger reliance on lake water as primary water source (70% of households in Kangatosa ward), with more urban wards reporting higher use of tap water (Township (77%), and Kanamkemer (92%)) (Table S3). Fishing was the primary household source of income for Kangatosa Ward (75%), while small business was the primary source of income for all other wards. Livestock as the primary source of income was most common in Kerio ward (31%) (Table S3).

Median household size was 6 people (IQR 5, 8).

#### Prevalence of malaria as reported by PCR and RDT

Malaria prevalence was 21% as reported by RDT (95% CI [8.8%-42%]) and 21% by PCR (95% CI [11%-38%]). By both RDT and PCR, malaria prevalence was the highest among individuals 5-19 years old in both sub-counties (Figure 2A).

**Figure 1.**
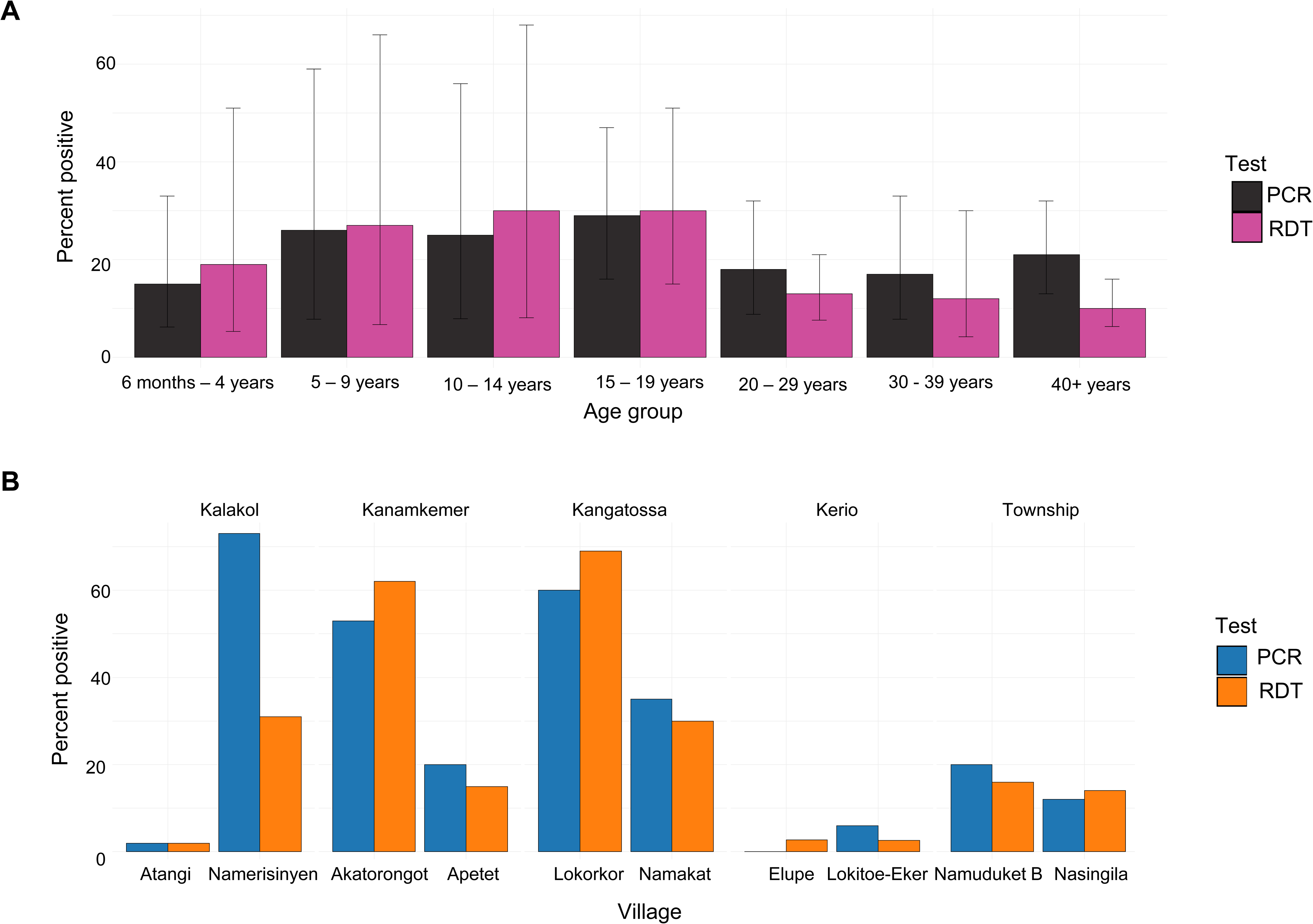
PCR and RDT positivity during quantitative baseline study, Turkana Central Sub-County, 2023. A. By age group, B. By Village.

**Figure 2.**
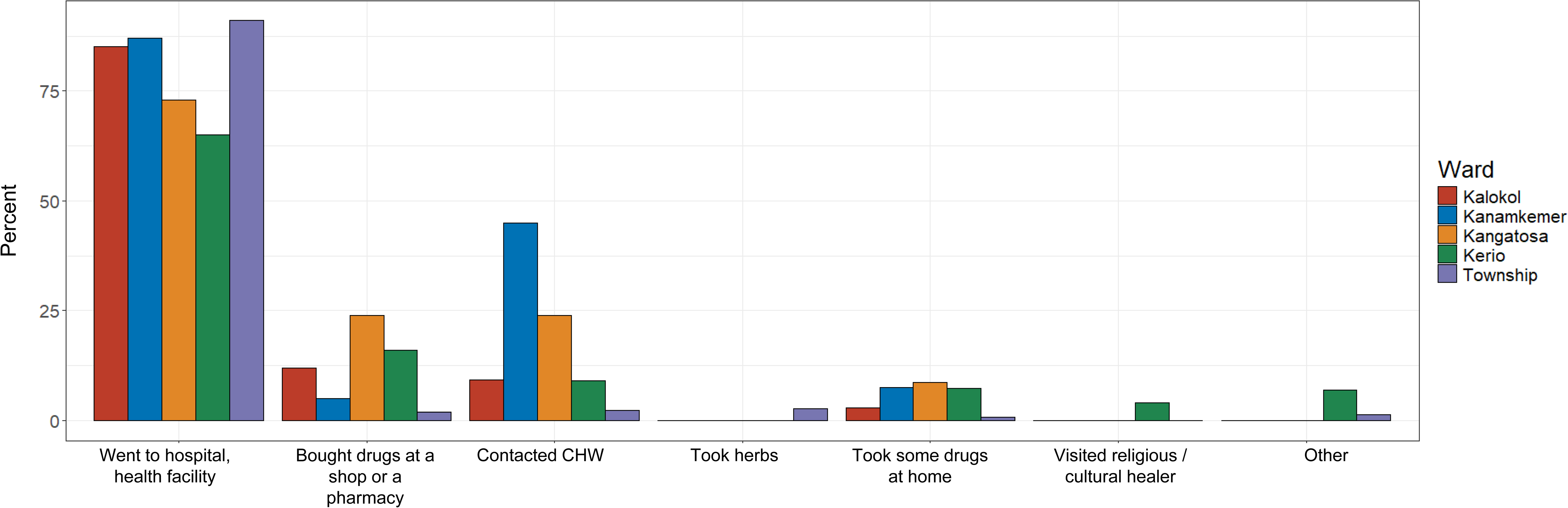
Actions reported when survey participants were asked “What did you do when you felt sick?”

Both RDT and PCR positivity varied widely across villages and wards. Kerio Ward had the lowest overall positivity using both PCR and RDT, followed by Township Ward (positivity <20% for both tests) (Figure 2B). However, in Kalokol, Kanamekemer, and Kangatosa, there was wider variability in results; in Kalokol Ward, by PCR, Atangi village had <2% positivity, while Namerisinyen had 73% positivity, although this difference was much smaller by RDT. In Akatorongot and Lokorkor villages, positivity was >50% both using PCR and RDT. While there was some variability in distribution of ages of individuals enrolled across villages and wards (Figure S1), this variability is unlikely to explain all of variability in PCR and RDT positivity.

46% and 62% of individuals with positive PCR and RDT results, respectively, did not report any malaria-like symptoms at the day of the survey (reported to be asymptomatic).

#### Knowledge and practices regarding malaria

Most individuals believed malaria was present in their area (98%); malaria to be a big problem in their village (98%), and that malaria can be prevented (94%) and cured through treatment (99%) (Table S4). While there was high perception that sleeping under a mosquito net protects against malaria, ownership and use of nets was very low as expected for a region where no mass distribution campaign has occurred. The proportion of households with a net for their sleeping space ranged from 20% in the most rural areas (Kerio Ward) to >50% in more urban township areas (Township and Kanamkemer Ward). Over one-fifth (22%) reported sleeping outside last night; this proportion was highest among individuals 40 years and older (37% [27%-50%]) and 15-19 years old (35% [22%-50%]), and the lowest among children under 5 years (11% [8.3%-13%]). However, amongst those who reported having a net available the night before the survey, 99% slept under it. Health education messages about malaria were often received from community health workers, with exception of Township ward, where radio was the most common source of information about malaria (47%) (Table S4).

#### Healthcare seeking

When asked what the individuals did when they experienced symptoms related to malaria, most reported that they went to the hospital / health facility, although this percentage ranged from 65% in Kerio Ward to 91% in Township Ward (Figure 3). In Kanamkemer Ward, almost half of respondents contacted a CHP indicating that CHPs are already a trusted resource for malaria in these areas.

### Results of the qualitative study

#### Profile of participants in qualitative study

Characteristics of individuals participating in FGDs, KIIs, and IDIs are presented in Table 2. In each ward, we enrolled at least ten leaders (five in each village). Profile of leaders enrolled was informed by interviews and understanding of the communities; for example, in Kalokol and Kangatossa wards, fishing is an important source of livelihood given proximity of the lake. As a result, in these wards, we carried out interviews with chairpersons of fisherfolk. In each ward, we interviewed at least two religious leaders (enrolling leaders of different religions when present), at least one chief or assistant chief, or another administrative leader in each of the villages (at least two per ward), leaders of women’s and youth groups, and other leaders depending on availability and relevance. In each ward, we carried out at least two interviews with health facility personnel or chairpersons of the health committee personnel. These were not done in villages where there was no health facility and no active health committee.

**Table 2.**
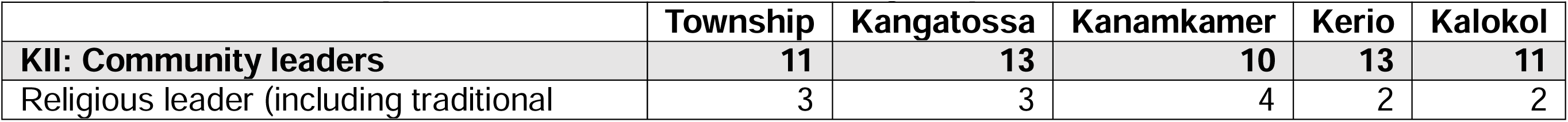
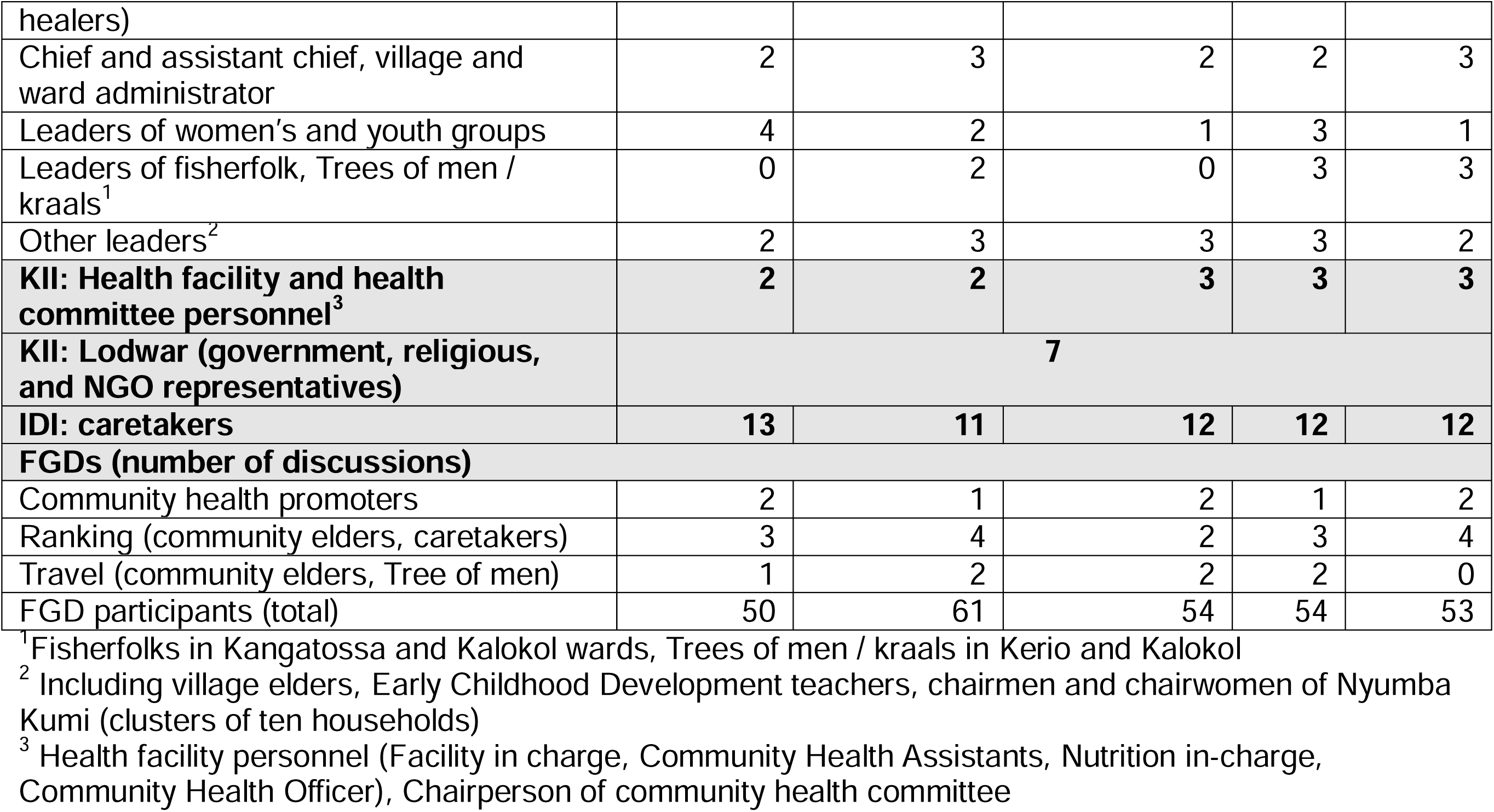
Profile of participants in the qualitative study, by ward. KII: Key informant interview; IDI: In-depth interview; FGD: Focus group discussion

Seven representatives of government, religious, and non-governmental organizations were interviewed. Furthermore, sixty caretakers participated in IDIs (11-13 per ward). Finally, at least two ranking FGDs were done in each ward; participants included caretakers of children under 5, or community elders. In areas with large amount of seasonal movement reported, travel FGDs were carried out either with community members (Township and Kerio wards) or members of the Tree of Men (Kangatossa, Kanamkemer, Kerio wards) (Table 2).

### Perception of SMC and potential challenges to uptake

The results are presented by the larger domain that could present challenges to reaching high SMC coverage, namely acceptability and reach. In each domain, we present findings organized by the relevant themes of the Designing for Behavior Change Framework, as presented in Table 3.

**Table 3.** Community perception of Seasonal Malaria Chemoprevention (SMC), by determinant of the Designing for Behavior Change Framework. Determinants are further classified into two domains that could present challenges to reach and uptake of SMC in Turkana Central Sub-County.

| Domain | Determinant | Description | Recommended action in SMC roll-out |
| --- | --- | --- | --- |
| Acceptability | Perceived severity | Malaria perceived as severe | No particular action necessary |
|  | Perceived susceptibility / risk | Children (and all other groups) overwhelmingly seen as susceptible to malaria | Coupling SMC with other interventions that can provide protection for other groups not targeted by SMC (e.g., elderly, pregnant people) |
|  | Perceived action efficacy | Perceived limited impact of SMC due to restriction in age group targeted and duration of SMC | Accompany SMC by distribution of mosquito nets and/or education activities on malaria prevention |
|  | Perceived positive consequences | Perceived reduction of morbidity and mortality among children; reduced financial burden due to costs of treatment of malaria | No particular action raised as many perceived positive consequences |
|  |  | Concerns raised about SMC being administered by anybody other than skilled health professionals | Sufficient, tailored training on administration of SMC to Community Health Promoters (CHPs) and caretakers of children under five years old |
|  | Perceived social norms | Support for SMC from neighbors, clans, and community leaders is crucial in increasing uptake among community members | Engaging community leaders (traditional, administrative, and religious) in mobilization, modeling behavior (serving as example) |
|  | Culture | Following structures were highly influential in decision-making, including around children's health, and in larger community life:<br>Religious structures<br>Nyumba Kumi (groups of ten households)<br>Kraals (groups of pastoralists moving together), and Tree of Men (larger groups of pastoralists sharing in decision-making)<br>Community groups (youth, business, women) | Engage in sensitization and mobilization within these structures, especially engaging leaders of these groups |
|  | Perceived negative | Few noted, but potential for misinformation | Concerted sensitization efforts |
|  | consequences | and growing fear of drugs, especially following COVID-19 vaccination campaign | focusing on content of drugs, dosing, potential side effects, and safety monitoring by community elders and other community structures |
| Reach | Access | Distance (pastoralists, sparsely populated communities, populations displaced due to conflict and flooding) | Outreach, use of CHPs (with transportation provided) |
|  |  | Individuals experiencing stigma (alcohol use, people living with HIV, children with disabilities) | Engaging community leaders to improve CHP access to households<br>resisting access<br>Sensitization and mobilization |
|  |  | Caretaker availability (caretakers working outside home in urban areas, farmers and fisherpeople in rural areas) | Sensitization and mobilization<br>Distribution on weekends and through centralized distribution points |
|  |  | School going children (children 3 – 4 years enrolled in Early Childhood Development centers would be missed if distribution were to happen on weekday mornings during school year) | Distribution on weekends and through centralized distribution points<br>Distribution in schools with parental consent |
|  |  | Perceived lack of access due to corruption of individuals involved in distribution | Engage religious leaders and women |

#### Domain 1: Acceptability

Community and individual acceptance of SMC will depend on the risk/concern for malaria, perceived benefit, and trust with groups implementing SMC.

##### Perceived severity

Overall, malaria had a high *perceived severity* as a disease that could result in death, with some community members believing it was responsible for many deaths.

In all KIIs with healthcare workers, malaria was named as a key health issue for the community, followed by respiratory infections, diarrhea, cholera and other waterborne diseases.

##### Perceived susceptibility/risk

The targeted population for SMC, namely young children under 5 years old, was seen as consistently susceptible to malaria. However, the *perceived susceptibility/risk* was not exclusive to this group. There were requests that SMC be expanded to include other population groups, especially the elderly and pregnant people, or that groups not covered by SMC be offered alternative strategies for malaria prevention, such as mosquito nets and repellants.

> *“What about us? Even if [children’s] immunity is low, malaria attacks even adults like us. Why can’t the government say that even adults should be given [SMC] for prevention, even though we are closer to death?” – Community leader, Township*

##### Perceived action efficacy

The limited population targeted (children under 5 years old) impacted the *perceived action efficacy* of SMC. In addition, in the Kangatossa ward especially, where Lake Turkana plays a vital role in the livelihood of community members, participants expressed concerns that malaria in their area is not limited to a single season but is present throughout the year.

> *“We will also need [SMC] during the other time because we are affected by malaria even when it is not a rainy season. We are neighboring a river, so there are mosquitos like all the time both during rainy season and during the dry season.” – Community leader, Kerio*

##### Perceived positive consequences

In general, SMC overwhelmingly had *perceived positive consequences* by all participants. These included improved health outcomes and reduction of mortality from malaria.

> *“It will help the children. It will help them so much. This issue of many deaths will reduce because of late we have been losing a lot of children to malaria. You can give birth to a child and [it] will even die within one day. […] This SMC will help reduce the number of child mortality because of malaria.” – Caretaker, Township*

In addition, participants mentioned that SMC would reduce financial pressure on caretakers of children because of averting malaria, which could further reduce intra-household tensions. The latter advantage was raised often in rural communities outside the Township ward, with participants noting that this would be of particular benefit to less wealthy households, where disposable income was scarce. One participant noted,

> *“I think [SMC] is good because for some people when their child gets malaria, they don’t have the money to take them to hospital. The child just suffers at home and sometimes they die.” – Community leader, Kalokol*

Furthermore, participants also reported that SMC would help children whose caretakers may not seek care at health facilities for reasons other than cost, such as households with a preference for traditional treatment.

Previous Community experience with other mass events, like health campaigns, may ease the acceptability of SMC. For example, the success of campaigns such as polio vaccination may contribute to the positive perception of SMC.

> *“I hope this activity will be good […], it is like the immunization of polio, because most of our children have been dying of polio, but when the immunization started, at least we are seeing it has reduced. People are not more lame now as they have been. I think for [SMC] also, although we have not seen it work.” – Kalokol, community leader*

Some caretakers described negative experiences with mass events. For example, one caretaker took time off work to attend food distribution; upon arrival at the distribution sites, they found a long queue of individuals that they did not feel met the eligibility criteria for distribution. Despite the wait, the caretaker did not receive food nor information about future distributions, and felt confused, frustrated, and reticent to attend future mass events, especially those organized by village elders.

CHPs were very highly regarded and trusted in most communities. Despite general positive perception of SMC’s potential, some participants raised questions about SMC being administered by anybody other than qualified medical personnel. Specifically, some participants expressed distrust of the CHP’s ability to provide a correct dose of SMC to the child, stating that they would only accept SMC from nurses or doctors.

##### Perceived social norms and culture

In addition to the primary caretaker’s acceptability of SMC, participants highlighted the importance of interpersonal- and community-level acceptability of SMC, with *perceived social norms* and *culture* playing a role in the caretaker’s decision on whether their child should take the drugs. Most commonly, when alive, the child’s mother was the primary decision-maker about health. However, many individuals influenced the mother, including neighbors, family members, community elders, CHPs, religious structures, and clans.

Community leaders often set and modeled social norms. Their engagement and acceptance of activities were consistently underscored as a major factor in their success and a requirement for the implementation of SMC. In addition, households reported being connected through the many religious groups in the area, with leaders playing an important role in healthcare decision-making and beyond. In addition to religious leaders, administrative leaders like area chiefs and ward and village administrators have considerable presence and influence in the communities. Together with village elders, these leaders were reported to be the major source of information across all wards.

Community leaders provided support by organizing barazas (community gatherings summoned by the chief or village elders) or sharing information about an event and sometimes serving as an example by modeling the desired behavior:

> *“My role, one is to mobilize the people, two is to help them have confidence in the activity that is supposed to be undertaken, because sometimes the people might be asking, why, why; like that one of Covid-19 [vaccination]. People had many misconceptions. They said, this one will kill, this one will do this. So, you give them the confidence and then you become the first person to be vaccinated, and then they wait for you. They say, let’s see if you can die. So, to create trust in them and the confidence, at least you must be the lead.” – Community leader, Kalokol*

Beyond formal leadership and religious leaders, communities had structures that linked community members based on livelihood activities or individual characteristics. These included youth groups, fisherfolk groups, women groups, table banking, and others, reinforcing connections between households and individuals. Regardless of the groups’ objectives, these also served as a mode of sharing information about community matters, including health. Additional community structures included the Nyumba Kumi, or clusters of ten households initially set up as a community policing concept [30] that has since extended to implementation of COVID-19 counter measures among others [31], and kraals among the pastoralists. A positive or a negative view of SMC by leaders and other members of these groups was likely to influence their members’ view and uptake of SMC.

##### Perceived negative consequences

There were few *perceived negative consequences* of SMC, although participants did raise questions about the contents of the drug, potential side effects, and risk of overdose to the child. In all wards, participants noted a possibility of misinformation contributing to perceived negative consequences of SMC, especially among community members with lower education levels. In a few communities, leaders noted an increased resistance towards medication/drugs; this resistance may extend to SMC. Participants often requested that SMC be accompanied by food and drinks for the children during administration to ensure they were not taking it on an empty stomach.

### Domain 2: Reach

In this domain, we present challenges classified under the *Access* determinant, namely hard-to-reach communities, households, and individuals; mobile population; individuals experiencing stigma; and other groups.

#### Hard-to-reach communities, households, and individuals

In general, *access* was the most reported challenge to the implementation of SMC. First, there are physical components to access that may impact SMC coverage. The geography poses physical barriers, such as rivers and mountains, limiting access for groups outside the Township and Kanamkemer wards. In sparsely populated Turkana Central Sub-County, individuals who live in far, difficult-to-access areas, such as interior rural areas and pastoralist communities, were often identified as at risk of missing SMC. Another group was those with less wealth and limited transportation options. In addition, there are multiple populations displaced by conflict, such as communities in Kerio, who regularly lack access to health facilities and CHPs due to the remote location of their new settlements. In at least one case, following conflict over cattle, a portion of the population relocated across the river from their original settlement and the health facility, with a small group settling in the mountains where they felt less at risk of future attacks. Strategies to reach this population include outreach and, to a smaller extent, distribution of SMC at central locations in the village during large market dates.

In addition to physical barriers to access, there are *access* concerns for school-going children. In Kenya, most children three to five years old attend Early Childhood Development and Education centers for pre-primary education [32], with some children attending madrasa, Islam-centered schools that fall outside the formal education sector. As a result, if SMC is administered while school is in session, many children in the target group would be absent from households during the day. Some participants suggested the distribution of SMC through schools by health workers, complemented by community-level or health facility distribution for other children. However, parental consent would have to be obtained before school-level administration, and children’s age eligibility would have to be confirmed. In madrasa schools, in particular, parents may feel strongly about the need to be present during administration, as reported by a religious leader.

On the other hand, children whose caretakers are unavailable during the workday would not receive SMC unless someone could provide approval. This was a particular problem in urban communities, especially in Township ward, where many primary caretakers leave home early in the morning and return late at night during the working week. Door-to-door strategies, or fixed-point distribution during workdays, would miss these children.

Although there exists a system of registering newly arrived households and individuals in households rosters, this process was reported to take up to six months. During this time, new arrivals may miss out on regular visits from CHPs and from being included in mass events, if the SMC campaign requested registration card prior to administration of SMC drugs.

Further, in some communities, participants reported perceived corruption of community elders or other individuals involved with mass events. Respondents suggested engaging women or religious leaders in distribution as a possible mitigation strategy, as the latter were perceived to be less corrupt.

#### Mobile populations

Travel and migration may mean that children’s access to healthcare is disrupted or changed. Although we identified some instances of short-term travel, such as visits to sick family members, temporary employment, and migration in the fisherfolk community, longer, predominantly seasonal migration by pastoralists is likely to pose a larger challenge to success of SMC. Specifically, we focused on understanding the movements, shift in healthcare seeking, and opportunities to reach pastoralist communities during migration.

Whether young children participated in pastoralist travel varied by location and household. It was most common that young and middle-aged men were the ones to migrate with the herd. These men would be accompanied by older children (usually about ten years and older), and women. Some households were divided in two, with the elderly, young children, people living with disabilities, and some women remaining behind, while others, usually smaller households, would migrate together. Children who attended school would often be left behind to continue their education. Some polygamist families would split, with one wife remaining behind, and others migrating. This complicated our understanding of how many children may actually leave their communities during migration, and would require additional efforts for mapping and quantifying how many of these children migrated during SMC.

Planning for reaching migrating children may also be affected by shifting and less stable migration periods. Until recently, drought seasons spanned about six months, with one dry season from January to April, and a shorter dry season from August to September. However, participants noted that recent shifts in climate made seasonality much less predictable. As a result, pastoralist migration occurs more sporadically, with pastoralists leaving in search of pasture whenever there is a drought. Pastoralists would return to their residence communities under three circumstances: when rains occurred, the pasture in migration destination was depleted, or they encountered insecurity. As SMC is likely to be administered just before and during rainy season, we anticipate that most of the monthly cycles would find pastoralists in their home locations; however, unstable rain patterns may result in some children migrating outside their communities during SMC administration.

Linking children to care during migration may be difficult. Often participants reported that travel would take place to an area far from a populated place and lack access to formal health system, except through rare outreach events, organized as mobile clinics brining services to these communities. Although CHPs played an important role in linking community to care, often providing screening and curative services, these connections are disrupted during migration. While some pastoralists reported that they were able to maintain contact with their CHPs by mobile phone, for others, there was no contact. Participants described that they often introduced themselves to the elders in the new location they migrated to, and were then linked to this community’s CHPs.

However, even after being assigned a new CHP, they reported receiving limited services. More often, pastoralists settled in an area outside of formal settlements, and their relationship with the new community was limited. As a result, pastoralists reported that their children often missed vaccination during migration.

Rarely, pastoralists reported returning to their residence communities to seek healthcare. However, participants stated that if they received information about a mass event like a vaccination campaign or SMC, they would likely return to their residence community. Often, they recommended using community leaders (as below), or CHPs, to provide this information by mobile phone.

Migration decisions were dynamic, and led by a group of initiated elders called the Tree of Men (Ekitoe a ng’ikiliok). While the quick initiation of migration may make it difficult to adapt SMC strategies to reach pastoralist children, existing relationships and communication strategies between the elders and the Tree of Men could be leveraged to facilitate service delivery during migration.

In addition to children who migrated with the pastoralists, we explored whether children who remained in their residence communities would experience additional barriers to SMC. Responds assured us that when the head of the household migrates, the remaining caretaker can make decisions about healthcare. While a young child usually remains with their mother if possible, extended family members or neighbors sometimes served as primary caretakers. In these cases, the caretaker may consult with elders, Nyumba Kumi (organization of ten households), CHPs, and other community members when making the decision about the child’s health. The father may become involved only when the child’s care requires financial resources outside the remaining caretaker’s ability to manage and hence would not impact SMC.

#### Individuals experiencing stigma

Stigma is an additional barrier to SMC from the perspective of both healthcare providers, including CHPs, and caretakers. There are caretaking groups, such as those struggling with alcohol use, that are negatively perceived by many healthcare providers, community leaders, and other community members. Several respondents mentioned that individuals struggling with alcohol use often caused disturbances, were not receptive to information about child’s health, and were unable to care for their children. When carrying out their activities, CHPs reported sometimes skipping households that struggled with alcohol use, while others recounted making several attempts to visit the household when the caretaker was not under the influence of alcohol. Children living in these households were reported to have been missed in both door-to-door and central location mass events.

There are two additional groups of children, those with disabilities and those living in households with a person living with HIV, that were identified as vulnerable to being missed. Children with physical disabilities, or those with caretakers with limited physical abilities, were often unable to attend distributions outside of the home, especially in rural areas. Some caretakers were reported to hide children with physical or mental disabilities due to shame and stigma and refused to provide access to their children to healthcare personnel, including CHPs. These children had lower access to and uptake of routine preventive and curative health services.

> *“Some mothers especially with disabled children tend to hide these children from CHVs [CHPs] and other community members, so they end up unattended.” – Community leader, Kerio*

While CHPs did not report negative attitudes towards children with disabilities, one community leader reported that some CHPs feel reluctant to work with these children due to the perception that some are dirty. For children with disabilities, using CHPs to deliver SMC at home was the predominant strategy suggested. However, the involvement of community leaders may be necessary to reduce the reluctance of caretakers to allow people in their homes.

In addition, CHPs reported that some people living with HIV were fearful that their status might be disclosed and, as a result, preferred not to allow CHPs or healthcare personnel into their households, no matter the reason for the visit. Some CHPs said they expected to be met with violence when visiting households with people living with HIV, resulting in a reluctance to visit. For example, when asked about households that were difficult for CHPs to work with, one CHP replied,

> *“That household where a person is HIV positive, whenever they see you, they will just start abusing you recklessly without any reason.” – CHP, Kalokol*

Community leaders, on the other hand, reported that people living with HIV were less likely to seek services at health facilities, where they were fearful of being systematically tested for HIV. Ensuring clear communication about the objectives of SMC and decoupling it from services that target people living with HIV may reduce the fear of stigma and disclosure.

Finally, children of sex workers were reported to less likely present to health facilities due to their caretakers’ fear of being systematically tested for HIV. However, participants did not report significant barriers to access of these children by CHPs in their homes or in locations outside health facilities.

### Approaches to ensuring high coverage with SMC

Despite the identified barriers to acceptability and access to SMC, participants identified opportunities and facilitators that can guide the design of the program strategy.

#### Sensitization and mobilization are crucial for success of SMC

In most interviews and discussions, participants highlighted the importance of community sensitization and mobilization prior to administering SMC. These were seen as necessary for acceptance and uptake of the event, and **participants linked insufficient sensitization to negative experiences with previous mass events**, contributing to vaccine hesitancy during the COVID-19 campaign and physical scuffles during relief food distribution.

Leaders often noted their desire to be involved in the activities from the early stages of design to the end of the project. Some had felt caught unaware by earlier mass activities done without consultation. Community members also expressed the desire to see community leaders’ buy-in. Elders were identified as leaders with close ties to the community and were often suggested as focal points to ensure accountability during the roll-out of SMC and report any issues afterward. Notably, some leaders expected remuneration as they engaged in activities.

Across all wards, participants reported receiving information through the chief’s barazas (organized discussions with community members), through village elders, CHPs and health workers, church, door-to-door, and through word of mouth, primarily through friends and neighbors. Less often, participants received information through community groups like women’s groups, WhatsApp, radio and public announcements, pamphlets and posters, and loudspeakers. Sharing information by phone has become more common over time, which is particularly important to households who live far away in less densely populated areas. These findings highlighted the importance of engaging multiple communication channels when sharing information and mobilizing the community before SMC. Information shared should address concerns raised by community members that led to *perceived negative consequences*. Additionally, respondents asked other questions (Appendix 2); preemptively addressing these during community sensitization may reduce confusion and frustration by community members.

When possible, SMC should be accompanied by the distribution of mosquito nets, as well as education activities on ways to reduce mosquito habitats and prevent malaria.

Less commonly, participants requested mosquito tents and repellents, although these are not items that have previously been distributed through government initiatives. The preference for coupling SMC with other interventions, such as providing food relief or improving water, sanitation, and hygiene, was widespread in rural areas and among pastoralists.

#### Modes of SMC delivery

In most ranking FGDs, there was not a single strategy for SMC delivery that was preferred by all participants. Rather, combination approaches for rolling out SMC were suggested, where a dominant strategy was expected to reach most eligible children, supplemented by an additional approach for hard-to-reach populations. Strategies, including their strengths and limitations, are described in Table S6. In urban and peri-urban wards, distribution through a central location, namely a health facility, was the <u>dominant strategy</u>. However, this approach is likely to miss children as detailed above and the perception that all services at health facilities require payment, also discouraged some participants from selecting this as a dominant strategy.

Supplementary strategies to reach these children included door-to-door, use of CHPs, use of a central location in the village for distribution, and distribution in schools.

Distribution through stages, or transportation hubs, was suggested to reach mobile populations in urban areas. Distribution through churches on Sunday was suggested to reach well-to-do households that were often resistant or inaccessible to CHPs.

In more rural wards, decentralized approaches for distribution were generally selected as <u>dominant strategies</u>, including door-to-door, CHPs, and distribution through churches. <u>Supplementary approaches</u> like outreach to improve access for pastoralists and children who travel with caretakers to the river and the lake for fishing were suggested. Central location distribution strategies and distribution through health facilities were also identified as supplementary strategies. Engaging with the elders and Tree of men would also be necessary for reaching pastoralist children.

While CHPs were seen as a crucial component of SMC rollout across all wards, whether by distributing SMC directly, identifying households with children under five years old, or as a part of sensitization and mobilization efforts, serious concerns were raised about support necessary to be provided to the CHPs. CHPs often reported being overwhelmed, with one CHP being responsible for as many as 145 households. Both CHPs and key informants reported competing priorities, and the need for support including transport, water, and increased stipends, in addition to reporting tools and operational supplies like bags and airtime.

## Discussion

In this study, we found high prevalence of malaria in Turkana Central Sub-County, with 15% positivity by PCR among young children 6 months – 4 years old. With only a quarter of individuals reporting sleeping under a mosquito net the night before the survey, and highly seasonal pattern of malaria cases, strategies like SMC may provide a feasible way to mitigate the burden of malaria in this area. Overall, we found strong support for SMC from community members, including caretakers of young children, community leaders, CHPs, and healthcare personnel. While SMC was largely acceptable, access is likely to be a significant barrier to reaching high coverage.

Physical access was challenging in rural and more remote communities due to distance between households, poor or absent roads, and challenging terrain. In urban communities, while households were relatively easy to reach, many caretakers were absent during the day or were inaccessible to CHPs. These findings are consistent with challenges to access documented in previous SMC distribution campaigns in the Sahel, especially in rural areas [33].

The pastoralist population in Turkana Central Sub-County is likely to face unique challenges in accessing SMC owing in part to changing climate patterns, leading to less predictable migration, and disruption of healthcare access during migration. In this study, participants identified strategies such as outreach and engagement with pastoralist leadership structures, including the Tree of Men, as a feasible approach to reach the pastoralists. Strategies like the One Health strategy, launched in Turkana County in 2023 [34–36], can provide additional opportunities to reach pastoralists through existing collaborations between Ministries, especially if SMC is provided alongside veterinary or water services. Collaboration between various Ministries working with pastoralist populations could facilitate implementation of SMC; for example, information collected by the Ministry of Livestock during the registration of pastoralists can be used to map children during migration. Initiatives like the Community Disease Reporters, which engage pastoralists in animal health surveillance, can be replicated for SMC. Training SMC focal persons among pastoralists who move with the kraals to distribute SMC in those locations may also be advantageous. This is consistent with literature highlighting the importance of training community drug distributors (CDDs) from mobile populations in increasing uptake of mass drug administration for diseases like trachoma [22,37]. Similarly, appropriate treatment of malaria and use of nets was significantly higher in nomadic Fulani communities that had trained representatives than those without [38]. However, deploying pastoralist SMC distributors may require more extensive training and supervision than CHPs or healthcare workers to ensure the community trusts the focal persons’ abilities sufficiently.

Caretakers and community leaders preferred a combination of approaches for effective SMC distribution, varying across urban and rural settings. Optimal strategies may rely on a dominant approach supplemented by a complementary secondary distribution strategy informed by the specific access challenges of each community. In more rural, interior areas, a door-to-door approach by CHPs or specialized health personnel is likely to work well, supplemented by well-tailored outreaches to reach pastoralists and people further in the interior. In these rural areas, static distribution through centralized locations like churches and health facilities would provide an alternative for those missed at home or through outreach. In urban areas, door to door efforts may miss those who work outside the home or who prefer to choose their healthcare provider. In this setting, SMC distribution at health facilities may supplement door-to-door or CHP distribution efforts.

In Turkana Central Sub-County, engaging with community leadership structures is crucial to SMC’s success. Leadership structures include established government structures (chiefs, village elders) and religious leaders, but also unique locally instituted networks such as Tree of Men, kraal leaders and irrigation committees. These networks not only have unique insight into community dynamics and broad reach to remote and mobile households, but could also provide accountability of SMC program to the community. Participants highlighted the powerful role that leaders could play as influencers and role models, particularly for new initiatives. Engaging existing leadership structures in planning, sensitization, distribution, monitoring, and follow-up is critical for the campaign’s success.

This study has several limitations. While during qualitative data collection, we made efforts to capture the experiences of individuals at different points of the power spectrum, we heavily relied on information from community leaders and CHPs to understand the community structure and identify individuals for qualitative data collection. It is likely that we missed individuals who were excluded from community participation. We were similarly unable to engage with pastoralists and individuals living on the other side of the mountains. This was especially the case in Kerio ward. While our teams made an effort to interview some pastoralists who came from faraway locations to the village on market days and to understand the particular challenges of this community from the Tree of Men and other community leaders, we were unable to obtain this information directly.

In the quantitative study, the major limitation was availability of up-to-date sampling frame. For the first stage of sampling, we relied on the list of villages and populations provided by the Ministry of Health; however, it is possible that some of the remote villages with little access to CHPs and health facilities were excluded from this frame. Furthermore, older children (>15 years old) and male adults were more often absent from households during time of data collection, as evidenced by larger proportion of children and female participants in the sample, compared to expected. Although our teams carried out mop-up to increase enrollment, the results are subject to selection bias. However, very few children under 5 years old were unavailable.

Despite these limitations, our findings showed that in Turkana Central Sub-County, SMC presents a promising approach for mitigating morbidity and mortality from malaria in a highly-vulnerable group. However, ensuring widespread SMC coverage requires a respectful and inclusive engagement with established community structures, and using context-tailored approaches including combination strategies for SMC delivery for hard-to-reach population. SMC offers a chance to integrate supplementary initiatives, such as distributing relief food, mosquito nets, and animal health programs. These identified strategies are transferrable to other contexts as SMC expands beyond the Sahel region.

## Data Availability

Data produced in the present study are available upon reasonable request to the authors.

## Acknowledgements

We are grateful to the administrative and traditional authorities of Turkana County, and Turkana Central Sub-County, for facilitating the implementation of this study. We are especially thankfully to the communities of the Turkana Central Sub-County for sharing their experience and perspectives with the study team, and for the Community Health Promoters, health facility personnel, and local authorities that facilitated access to the communities and logistical support for both the quantitative and qualitative components of the study. We are grateful to Chrisine Markwalter for her support with planning the logistics of data collection. Finally, we are grateful to the data collection team and the supervisors, as well as transcribers and translators, who tirelessly worked to collect and make available the best possible data, without whom this work would not have been possible.

## Financial support

This study was funded by the Catholic Relief Services.

## Conflict of interest

The team does not report any conflicts of interest.

## Author’s current addresses

NK: Johns Hopkins Bloomberg School of Public Health, Baltimore, USA; CK: Academic Model Providing Access to Health Care (AMPATH), Eldoret, Kenya; TJC: Academic Model Providing Access to Health Care (AMPATH), Eldoret, Kenya; GA: Academic Model Providing Access to Health Care (AMPATH), Eldoret, Kenya; LA: Academic Model Providing Access to Health Care (AMPATH), Eldoret, Kenya; BR: Academic Model Providing Access to Health Care (AMPATH), Eldoret, Kenya; JKK: Academic Model Providing Access to Health Care (AMPATH), Eldoret, Kenya; EK: Academic Model Providing Access to Health Care (AMPATH), Eldoret, Kenya; ER: Duke Global Health Institute, Duke University, Durham, USA; KL: Unaffiliated, Lodwar, Kenya; DE: Turkana County Ministry of Health, Lodwar, Kenya; GL: Turkana County Ministry of Health, Lodwar, Kenya; OO: Catholic Relief Services (CRS), Nairobi, Kenya; SVH: Catholic Relief Services (CRS), Nairobi, Kenya; RM: Catholic Relief Services (CRS), Nairobi, Kenya; WPOM: Duke Global Health Institute, Duke University, Durham, USA; AW: Johns Hopkins Bloomberg School of Public Health, Baltimore, USA; BG: Johns Hopkins Bloomberg School of Public Health, Baltimore, USA; DM: Moi University School of Public Health, Eldoret, Kenya

## Notes

### Competing Interest Statement

The authors have declared no competing interest.

### Funding Statement

This study was funded by Catholic Relief Services.

### Author Declarations

Research Ethics Committee of Moi University gave ethical approval for this work. Institutional Review Board of Duke University gave ethical approval for this work.

