## Supplementary materials for "Community Informed Approach for Designing Seasonal Malaria Chemoprevention Delivery in Semi-Nomadic Population in Turkana Central Sub-County, Kenya: Results from Baseline Mixed-Methods Study"

Table S1. Profile of participants and thematic areas for the qualitative component of study, Turkana Central Sub-County

| Approach | Participant profile | Thematic areas |
| --- | --- | --- |
| Focus Group Discussions | - Community elders - Tree of Men | Travel |
|  | - Community health promoters | Community Health Promoter structure and challenges |
|  | - Caretakers of children under 5 years old - Community elders | Ranking exercise |
| Key Informant Itnerviews | - Community leaders | Social and resource mapping |
|  | - Community leaders - Government, religious, and NGO representatives (Lodwar) | Previous mass events and SMC |
|  | - Heads of health facilities, Community Health mobilizers and Community Health Assistants - Community health committee members | Community health profile |
| In-Depth Interviews | - Caretakers of children under 5 years old | Child’s healthcare seeking |
|  |  | SMC and mass distribution events |

Appendix 1. Quantitative questionnaire

Questionnaires

### Household survey

**Household member details**:

1. Questionnaire date (DD/MM):
2. Household ID:

**Household questions:**

3. How many people are in your household (provide complete exact number)?
4. How many bednets for sleeping under does your household own?
5. Do you have enough bednets for all the sleeping spaces?
6. If not, what is the main reason for not having enough mosquito nets in your household?
A. Bed nets are not available in the area
B. The cost is too high
C. The available bed nets are not effective in preventing malaria
D. Don’t think it prevent malaria at all
E. We don’t like to use them
F. No malaria in the area, we don’t need them
G. Can’t hang them (sleep outside or can’t hang them in their housing structure)
H. Other ____________________
7. What is the primary water source at the homestead? Circle all that apply
   b. River
   c. Lake
   d. Dam or water pan
   e. Hand dug water pit or hole
   f. Spring
   g. Well or borehole
   h. Tap water
   i. Other: _______________
8. What kind of animals are in your herd now and how many? If none, write 0
a. Goats (specify number):
b. Sheep (specify number):
c. Camels (specify number):
d. Cattle (specify number):
e. Other (specify name and number):

**Income question:**

9. What is your household’s primary source of income? Circle all that apply
   b. Livestock
   c. Farming
   d. Relative working in town
   e. Small business
   f. Other (specify): ____________

### Household member Enrollment Survey

This enrollment survey is for all household members.

**Household member details**:

1. Questionnaire Date (DD/MM):
2. Household ID:
3. Household member ID:
4. Age in years (write complete age):
5. Gender (circle): a. Male	b. Female	c. Other
6. Relationship to head:
a. Head
b. Wife
c. Child of head
d. Parents of head
e. Sister/brother of head
f. Cousin/niece/nephew
g. Aunt/uncle
h. Other (specify): __________

**Malaria RDT questions**:

8. Malaria-like symptoms today? Circle all that apply:

a. None
b. Fever
c. Headache
d. Body aches
e. Chills
f. Sweats
g. Sore throat
h. Cough
i. Diarrhea
j. Nausea
k. Vomiting
l. Stuffy/runny nose
m. Trouble breathing
n. Other

**If symptoms are reported, answer questions 9 – 11; otherwise, skip to 11**.

9. If symptoms reported, how long ago did they start?
   a. < 1 day
   b. 1-2 days
   c. 3-4 days
   d. 5-6 days
   e. 1-2 weeks
   f. ≥ 3 weeks
10. Where were you when the symptoms began?
    a. At the homestead
    b. Visiting another homestead
    c. Traveling with herd
    d. In town (specify): ________________
    e. Other (specify): ________________
12. **RDT Results:** a. Positive	b. Negative	c. Invalid

**Malaria prevention**:

13. Did you sleep at this homestead last night? a. Yes	b. No
    a. Did you share a sleeping space with others (eg. same bed, under same net, in same room/area)? a. Yes	b. No
14. If yes, specify how many others were in the shared sleeping space: ________________
15. Did you sleep inside or outside of the house last night? a. Outside	b. Inside
16. Was there a net available last night? a. Yes	b. No

**If a net is available, answer questions 16-20; otherwise skip to 21**

17. Did you sleep under a bed net last night? a. Yes	b. No
18. Did you share a bed net last night? a. Yes	b. No
19. Where was the net obtained?
    b. Health facility
    c. Mass distribution/donation
    d. Bought it
    e. Other (specify): __________________
20. How long ago did you get it?
    b. < 1 year
    c. 1 – 2 years
    d. 3-5 years
    e. > 5 years
    f. Don’t know
21. Does the net have holes? *Ask to see the net*
    b. None – good condition
    c. Some smaller than a coin
    d. Some smaller than a hand
    e. Large tears, bigger than hand

**Medical history**:

22. Have you been sick in the last month? a. Yes	b. No

**If sick in the last month, answer questions 22 – 26; otherwise, skip to 27.**

23. Where were you when you started feeling sick most recently?
    b. At the homestead
    c. Traveling with herd
    d. Visiting another homestead
    e. In town (write in name): ______________
    f. Other: ________________
24. What did you do when you felt sick?
    b. Contact CHW
    c. Go to hospital, health facility
    d. Bought drugs at a shop or pharmacy
    e. Took some drugs at home
    f. Took herbs
    g. Visit religious/cultural healers
    h. Other: __________________
25. Did you have a blood test done for malaria? a. Yes	b. No	c. Not sure
26. If yes, was it positive or negative? a. Positive	b. Negative
27. Did you take any of these medicines, regardless of being tested?
    a. None
    b. Antimalarial
    c. Antibiotic
    d. Pain Killers
    e. Don’t know/don’t remember
    f. Other: __________________
28. If the answer to Q26 is “None”, what was the main reason for not taking treatment? Do not prompt, may select more than one answer.

A. Health facilities are not available in the area, too far to travel

B. There are no CHWs

C. The cost for treatment too high

D. The available medicine are not effective for curing the illness

E. There was no one to take Him/her for treatment

F. Don’t think it cures the illness

G. Other____________________

**Travel history**:

29. Have you traveled outside of the area and stayed overnight in the last two months (for work, to travel with herds, to visit family, for a funeral, attend boarding school)? a. Yes	b. No

**If yes, complete questions 28 – 29; otherwise, skip to 30.**

30. If you have traveled in the last 2 months, list all trips where you traveled outside of the area and stayed overnight. Start with the most recent:
    a. Trip 1:
       ii. Subcounty (write in):
       iii. Nearest Town (write in):
       iv. How long did you stay there?
           b. < 1 week
           c. 1-2 weeks
           d. 3-4 weeks
           e. 1 month or longer
       v. Purpose of trip:
          b. Visit family/ friends
          c. Business /work
          d. Travel with herd
          e. Attend boarding school
          f. Attend funeral
          g. Other: _____________
    b. Trip 2:
       ii. Subcounty (write in):
       iii. Nearest Town (write in):
       iv. How long did you stay there?
           b. < 1 week
           c. 1-2 weeks
           d. 3-4 weeks
           e. 1 month or longer
       vi. Purpose of trip:
           b. Visit family/ friends
           c. Business /work
           d. Travel with herd
           e. Attend boarding school
           f. Attend funeral
           g. Other: _____________
    c. Trip 3:
       ii. Subcounty (write in):
       iv. Nearest Town (write in):
       v. How long did you stay there?
          b. < 1 week
          c. 1-2 weeks
          d. 3-4 weeks
          e. 1 month or longer
       vii. Purpose of trip:
            b. Visit family/ friends
            c. Business/work
            d. Travel with herd
            e. Attend boarding school
            f. Attend funeral
            g. Other: _____________
31. If you have traveled in the last 2 months, have you been treated for malaria after returning from your last trip? 1. Yes	2. No

****QUESTIONS FOR RESPONDENTS OVER 18 YEARS ONLY****

32. Do you plan to remain at the homestead or travel with the herd on the next migration?
    a. Remain	b. Travel with herd	c. Other (specify): ____________________
33. Describe where you will go with the herd:

Location 1

i. Landmark (describe):
ii. When were you last there? (start and end dates: DD/MM – DD/MM):
iii. How long did you walk before you arrived at this location (specify hours or days)? ________
iv. How long did you stay there?
    a. < 1 week
    b. 1-2 weeks
    c. 3-4 weeks
    d. 1 month or longer
v. Group (circle): a. main camp	b. satellite camp
vi. Were there other nomads not from your household nearby (overnighting within 100 meters)?:	a. Yes	b. No
vii. Number of people at this camp, including household and non-household members:
     a. 1-3
     b. 4-6
     c. 7-10
     d. 11-15
     e. 15-20
     f. 20+
ix. Water source nearby for herd (circle all that apply):
    a. River
    b. Lake
    c. Dam or water pan
    d. Hand dug water pit or hole
    e. Spring
    f. Well or borehole
    g. None
    h. Other: _______________
x. Which livestock were present in your herd at this location? Circle all that apply:

a. Sheep, milking
b. Sheep, non-milking
c. Goats, milking
d. Goats, non-milking
e. Camels
f. Cattle
   3. Location 2
      i. Landmark (describe):
      ii. When were you there? (DD/MM):
      iii. How long did you walk before you arrived at this location (specify hours or days)? ________
      iv. How long did you stay there?
a. < 1 week
b. 1-2 weeks
c. 3-4 weeks
d. 1 month or longer
   v. Group (circle): a. main camp	b. satellite camp
   vi. Were there other nomads not from your household nearby (overnighting within 100 meters)?:	a. Yes	b. No
   vii. Number of people at this camp, including household and non-household members:
a. 1-3
b. 4-6
c. 7-10
d. 11-15
e. 15-20
f. 20+
   ix. Water source nearby for herd (circle all that apply):
a. River
b. Lake
c. Dam or water pan
d. Hand dug water pit or hole
e. Spring
f. Well or borehole
g. None
h. Other: _______________
   x. Which livestock were present in your herd at this location? Circle all that apply:
a. Sheep, milking
b. Sheep, non-milking
c. Goats, milking
d. Goats, non-milking
e. Camels
f. Cattle
   5. Location 3
      i. Landmark (describe):
      ii. When were you there? (DD/MM):
      iii. How long did you walk before you arrived at this location (specify hours or days)? ________
      iv. How long did you stay there?
a. < 1 week
b. 1-2 weeks
c. 3-4 weeks
d. 1 month or longer
   v. Group (circle): a. main camp	b. satellite camp
   vi. Were there other nomads not from your household nearby (overnighting within 100 meters)?:	a. Yes	b. No
   vii. Number of people at this camp, including household and non-household members:
a. 1-3
b. 4-6
c. 7-10
d. 11-15
e. 15-20
f. 20+
   ix. Water source nearby for herd (circle all that apply):
a. River
b. Lake
c. Dam or water pan
d. Hand dug water pit or hole
e. Spring
f. Well or borehole
g. None
h. Other: _______________
   x. Which livestock were present in your herd at this location? Circle all that apply:
a. Sheep, milking
b. Sheep, non-milking
c. Goats, milking
d. Goats, non-milking
e. Camels
f. Cattle

32.	Have you ever heard about malaria? If your answer is ‘No’ Skip to Q ‘43’

1.	Yes

2.	No

33.	Is there malaria in this area?

1.	Yes

2.	No

3.	Don’t know

34. Is malaria a big problem in your village?

1. Yes

2. No

3. Don’t know

34.	How does someone get malaria? NB ; circle that all apply, do not prompt

A.	Being near other people who are sick

B.	Rainy or cold season

C.	Drinking things that are too cold

D.	Flies

E.	Mosquito bites

H.	Other___________________________(specify)

I.	Don’t know

35.	How can you tell if a persons have malaria? NB: circle that all apply, without prompt

A.	Fever

B.	Sweating

C.	Headache

D.	Chills/shivering

E.	Poor appetite

F.	Vomiting

G.	Cough/breathing difficult

H.	Diarrhea

I.	Joint pain

J.	Convulsion

K.	Coma

L.	Other _______________(specify)

M.	Don’t know

36.	Do you believe malaria can be prevented? If your answer is ‘No or Don’t know’ skip to Q ‘40’

1	Yes

2	No

3	Don’t know

37.	How can you prevent yourself from getting malaria? NB; circle all that apply do not prompt

A.	Isolating infected person

B.	Avoiding movement or staying at home

C.	Sleeping under a bednet

D.	Spraying the house with chemicals

E.	Taking specific herbs or foods

F.	Taking drugs

G.	Avoiding pond and dams

H.	Clearing bushes and avoiding swampy areas

I.	Other________________________(specify)

38.	Have you ever used one of the activities you just mentioned for malaria prevention?

1.	Yes

2.	No

39.	Which activities have you ever used to prevent yourself from getting malaria? Circle all that apply, do not prompt

A.	Isolating infected person

B.	Avoiding movement or staying at home

C.	Sleeping under a bednet

D.	Spraying the house with chemicals

E.	Taking specific herbs or foods

F.	Taking drugs

G.	Avoiding pond and dams

H.	Clearing bushes and avoiding swampy areas

I.	Other_______________________(specify)

40.	Do you believe that malaria can be cured through treatment?

1.	Yes

2.	No

3.	Don’t know

41.	Have you ever heard of mosquito nets that can be used to cover your sleeping bed /place while sleeping? If your answer is ‘No’ skip to Q44

1.	Yes

2.	No

42.	Do you think sleeping under the mosquito net protects from malaria?

1.	Yes

2.	No

3.	Don’t know

43.	Who should sleep under a bednet? Skip to Q44

1.	Children

2.	Pregnant women

3.	Women

4.	Everyone

5.	Guests

6.	Household head

44. In the past six months, have you seen or heard any messages about malaria? If your answer is

‘No’, skip to 46.

45. Where did you see or hear these messages? Circle all that are mentioned

RADIO . . . . . . . . . . . . . . . . . . . . . . . . . . . . . . . . . . A

TELEVISION . . . . . . . . . . . . . . . . . . . . . . . . . . . . . . . B

POSTER/BILLBOARD . . . . . . . . . . . . . . . . . . . . . . . C

NEWSPAPER/MAGAZINE . . . . . . . . . . . . . . . . . . . . D

LEAFLET/BROCHURE . . . . . . . . . . . . . . . . . . . . . . . E

HEALTHCARE PROVIDER . . . . . . . . . . . . . . . . . . . . F

COMMUNITY HEALTH WORKER . . . . . . . . . . . G

SOCIAL MEDIA . . . . . . . . . . . . . . . . . . . . . . . . . . . . H

RELATIVE/FRIEND . . . . . . . . . . . . . . . . . . . . . . . I

COMMUNITY DIALOGUE / BARAZA . . . . . . . . . . . J

COMMUNITY LEADER / ELDER. . . . . . . . . . . . . . . . . K

COMMUNITY EVENT / ROADSHOW . . . . . . . . . . . L

SCHOOL PUPILS . . . . . . . . . . . . . . . . . . . . . . . . . M

ANYWHERE ELSE X________________

46. Now I am going to read some statements and I would like you to tell me whether you agree or disagree with it. If you don't know, say, don't know.

a. I am confident in my ability to hang a mosquito net in my household. Do you agree or disagree?
b. People in my community usually take their children to a health care provider on the same day or day after they develop a fever. Do you agree or disagree?
c. People in my community generally trust/follow the advice given at the health facility. Do you agree or disagree?
d. People in my community who have a mosquito net usually sleep under a mosquito net every night Do you agree or disagree?
e. When a child has a fever, you almost always worry it might be malaria. Do you agree or disagree?

**Table S2. Enrollment levels for individuals, quantitative baseline survey in Turkana Central and North Sub-Counties**

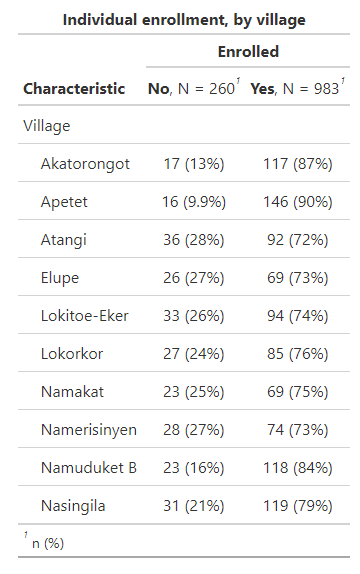

**Table S3. Household-level characteristics, Turkana Central and North Sub-Counties**

| **Characteristic** | **Kalokol** | **Kanamkemer** | **Kangatosa** | **Kerio** | **Township** |
| --- | --- | --- | --- | --- | --- |
| Primary water source |  |  |  |  |  |
| Tap water | 46% | 92% | 0% | 37% | 89% |
| River | 29% | 0% | 9.1% | 48% | 16% |
| Well or borehole | 23% | 4.4% | 7.3% | 16% | 5.9% |
| Dam or water pan | 0.9% | 3.3% | 0% | 0% | 4.1% |
| Lake | 8.6% | 0% | 70% | 6.5% | 0% |
| Hand dug water pit or hole | 3.0% | 0% | 22% | 4.2% | 0% |
| Spring | 0% | 0% | 0% | 0% | 0% |
| Other | 0% | 0% | 0% | 0% | 0% |
| Household primary source of income |  |  |  |  |  |
| Farming | 0% | 2.3% | 3.1% | 7.1% | 0% |
| Fishing | 3.1% | 0% | 75% | 0% | 0% |
| Livestock | 10.0% | 13% | 3.7% | 31% | 0% |
| Relative working in town | 11% | 12% | 0% | 8.7% | 15% |
| Small business | 66% | 69% | 22% | 50% | 72% |
| Other | 11% | 17% | 2.8% | 9.2% | 26% |

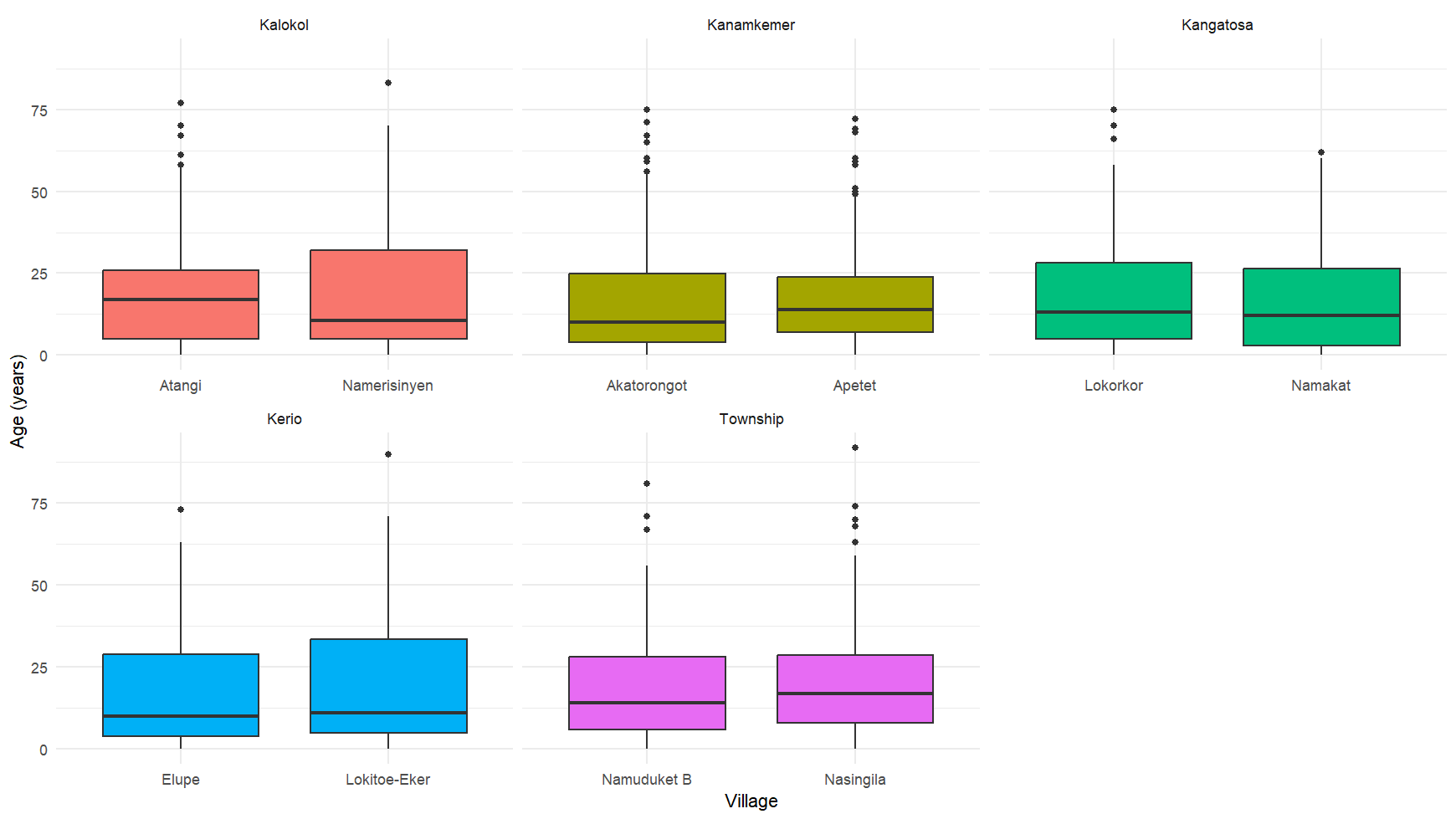

Figure S1. Distribution of ages in years for individuals enrolled in the survey, by village and ward

**Table S4. Individual-level malaria knowledge**

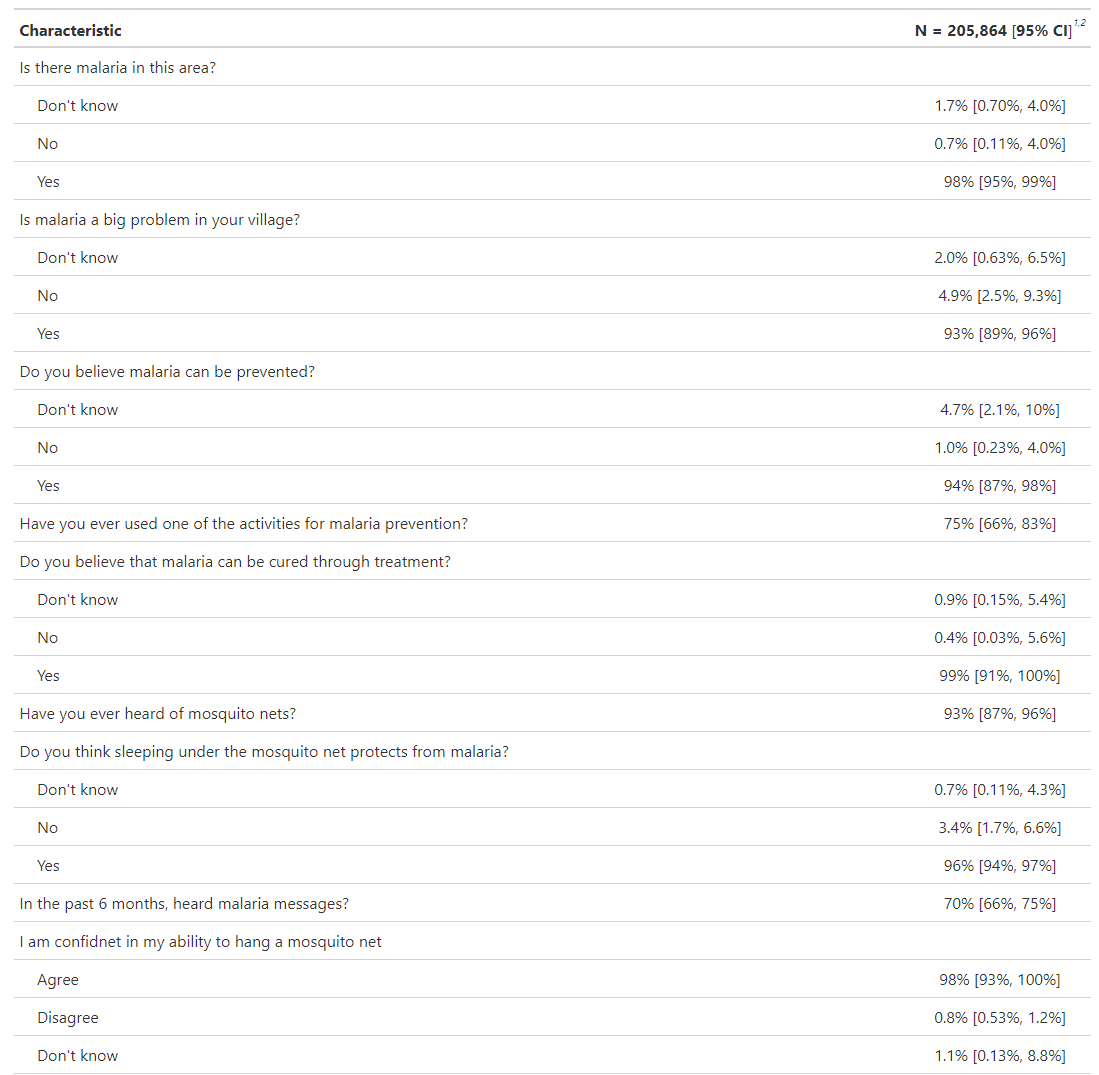

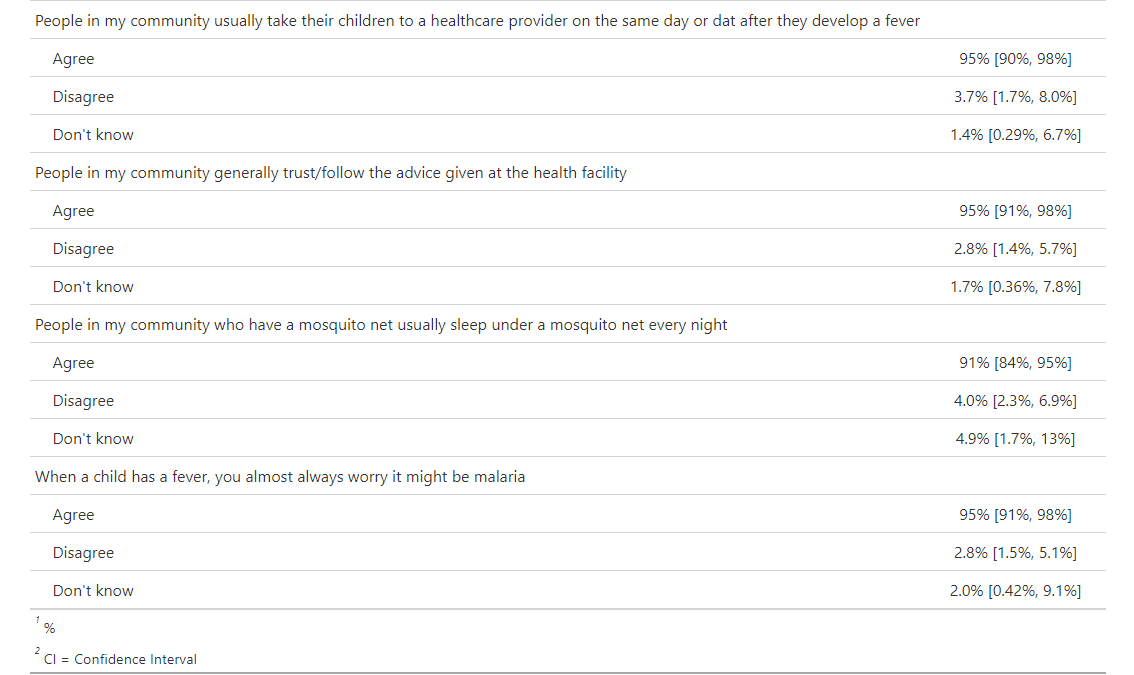

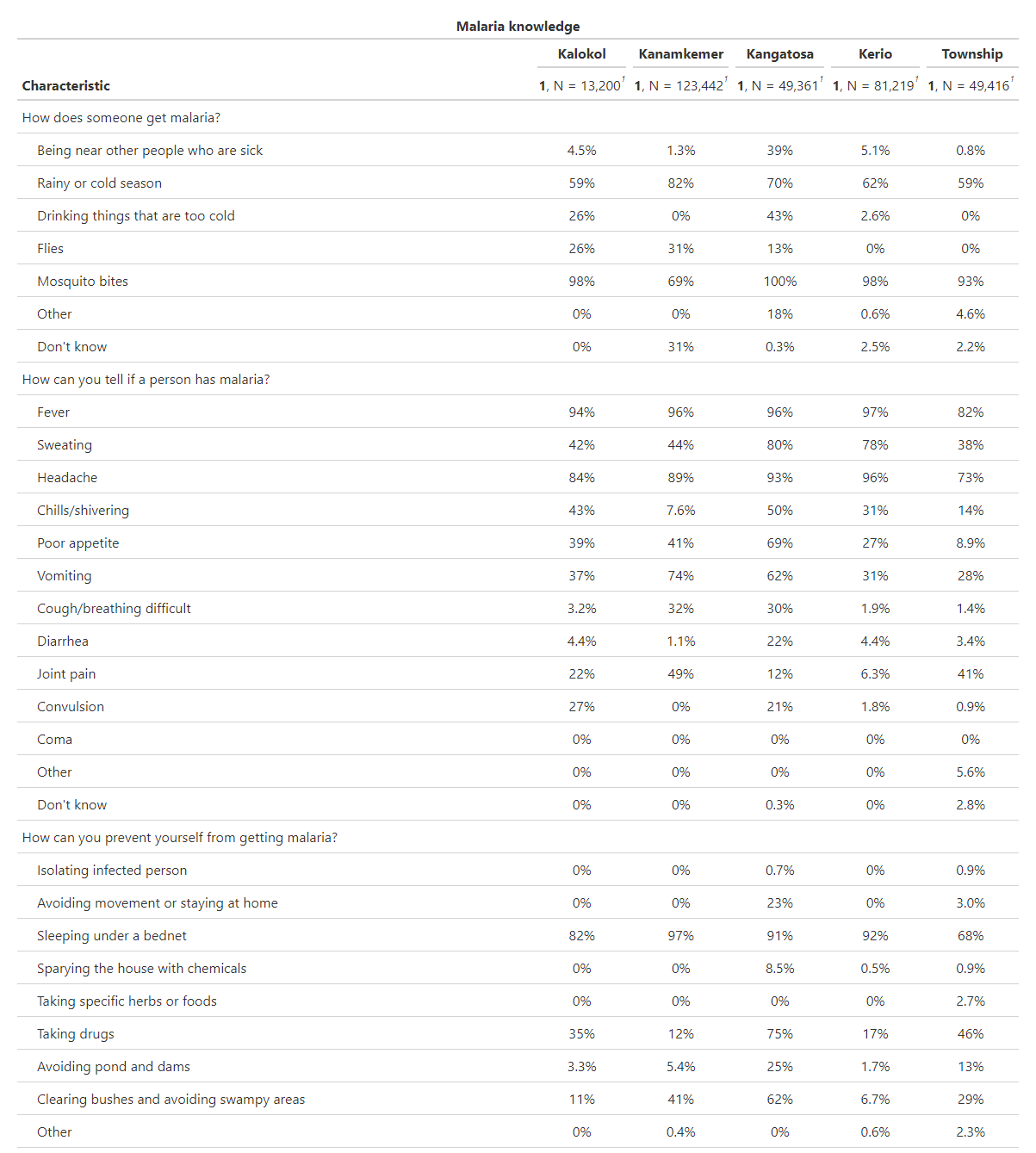

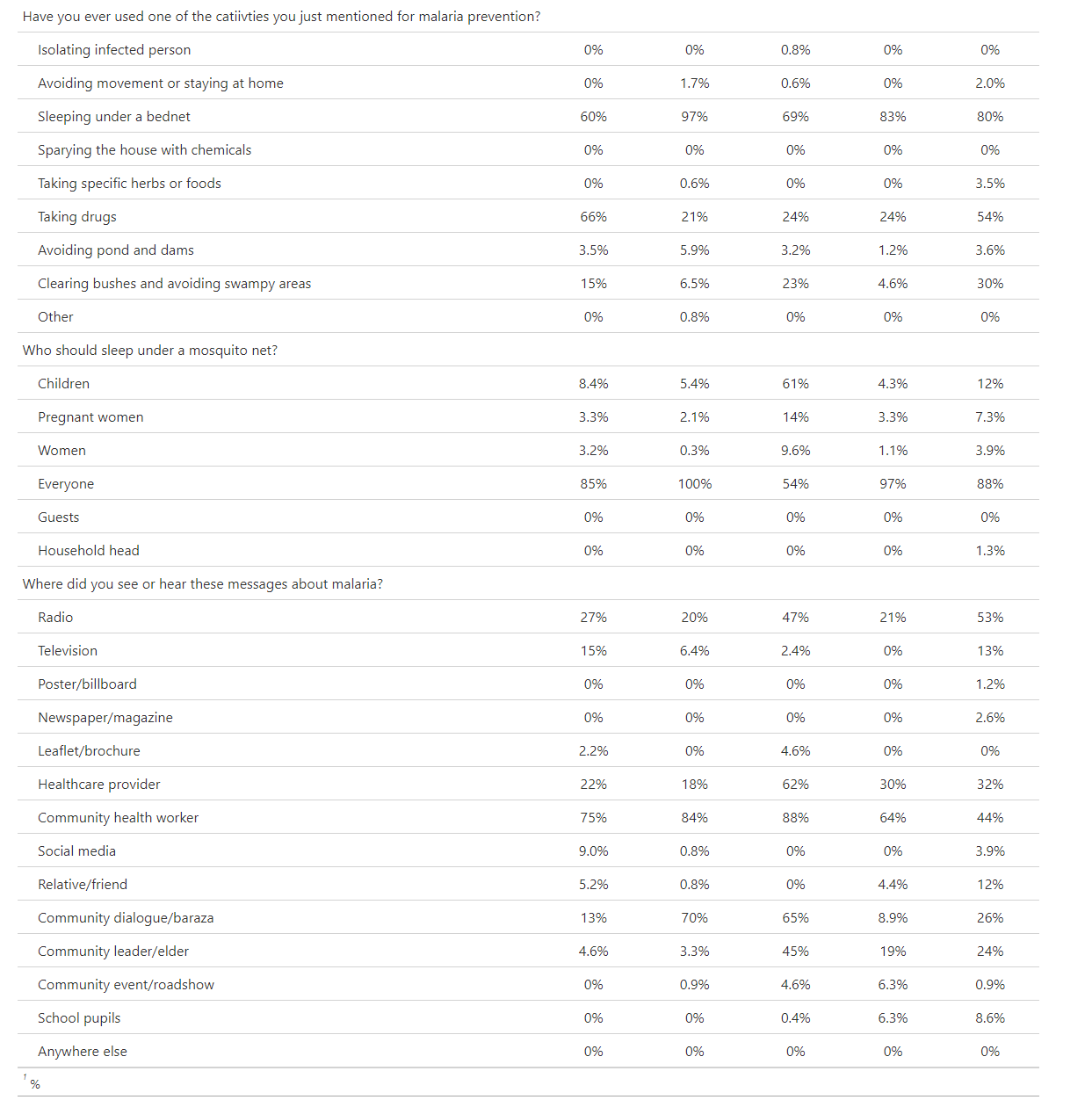

**Table S5. Malaria prevention activities (individual-level)**

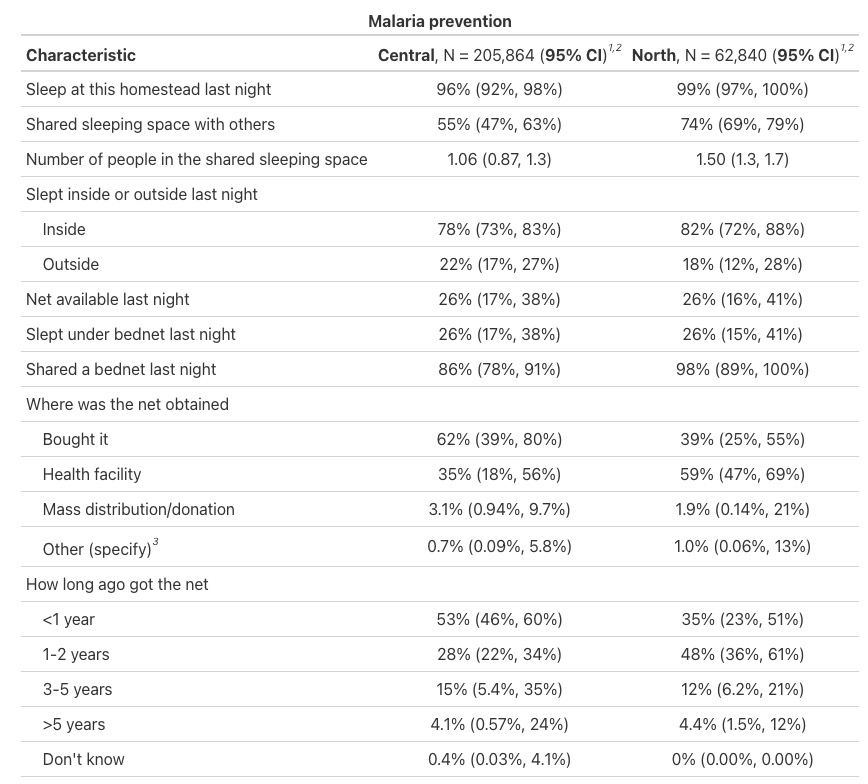

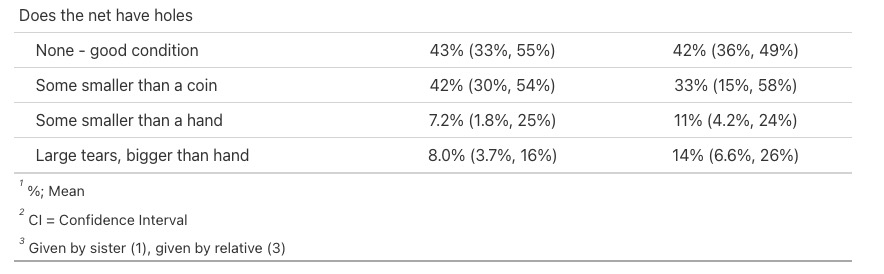

**Table S6. Strengths and limitations of possible approaches to administer SMC in Turkana Central Sub-County, identified through interviews and discussions with participants.**

| **Distribution approach** | **Description** | **Strengths** | **Limitations and Challenges** | **Most commonly preferred strategy** |
| --- | --- | --- | --- | --- |
| Health facility | SMC available through health facilities | Community is already familiar with seeking health services at the facility.  All drugs must pass through the health facility.  May facilitate seeking care for other health needs at the same time as getting SMC.  Availability of qualified personnel, increasing trust for surveilling and managing side effects.  Open at all times including night and weekends. | Long waiting times and overcrowding.  Some perception of corruption, and preferential treatment of clients.  Expectation that clients will have to pay for SMC like they do for other facility services.  Not all villages have a health facility nearby.  Caretakers have to bring children to the facility to get SMC. | X |
| CHPs | CHPs receive SMC and distribute it in the community. This may include door-to-door, at a specific location in the community, or having community members seek SMC from CHPs at their homes. | CHPs are already a known and trusted source of healthcare, especially in remote communities.  CHPs know the households and their composition.  Short distance to reach the CHPs as they live in the community.  CHPs can revisit households if missed.  Experience mobilizing communities.  Services provided by CHPs are free. | Lack of transport, difficult working conditions.  Grudges and lack of respect towards and from some households.  Competing activities.  Some distrust towards CHPs’ ability to distribute drug.  Require additional incentives and motivation, including higher financial incentives and logistical support. | X |
| Door-to-door | Nurses or other health personnel carrying out SMC door-to-door, engaging CHPs for identification of households with eligible children | No cost of transport or time sink for caregivers.  Removes barrier for children with disabilities.  Health personnel more trusted in administration of drugs and handling side effects.  Can be used to diagnose / screen for other conditions as well. | Unlikely to revisit households that are locked / where children or caregivers are absent.  Resource-intensive.  May have to work before and after workhours and on weekends.  Need support from CHPs to identify households with children. | X |
| Church | SMC distributed following Sunday service or Sunday school | Church leaders are highly influential.  Many people attend church, including those that would be missed during weekday distribution.  Perceived humility of church attendees on Sunday, including those that would refuse SMC elsewhere. | Orphan and disabled children may be missed, as well as children that live far.  Some children come to church without their primary caregivers.  Church services only on Sundays.  Miss people that don’t attend church; need to engage with all denominations and religions in the community. | X |
| School | SMC distributed by healthcare providers through Early Childhood Education schools | Large number of children 3 years and up attend school.  If not used for administration, the teacher could send the children to the health facility to receive SMC.  Teachers generally trusted to take care of the children; have experience with child illness when child falls ill at school. | Some children do not attend school (under 3, and some children that are eligible).  Caregivers must be present at the time of SMC administration, or provide consent prior to administration.  School not open the entire year, and sometimes there are school closures outside of academic calendar. |  |
| Outreach | Mobile clinic-type approach, where SMC is offered in a fixed location with hard-to-reach population, like farmers or pastoralists | Can reach children that would be missed otherwise, including kraals as they are on the move, children going with their mothers to the river, and children that are living far.  Strategy that’s already been a part of health system.  Potential to integrate in the OneHealth framework with other services. | Resource-intensive.  Hard to reach areas can still be very difficult to reach. | X |
| Chief camp / village admin / elders^1^ | Distribution by healthcare providers or CHPs at large meetings organized by community chief, administrators, and village elders | Highly influential people.  Community members come in large numbers when called.  These are known points of meetings, so are good as central mobilization points.  Meetings can be called as needed (e.g., on a monthly basis for SMC) | Young children typically do not attend these, unless specifically requested.  Usually for distribution of food or goods, which may upset people if they feel tricked.  Not everybody comes.  May be a better approach for mobilization of community for SMC rather than distribution. |  |
| Central distribution / market places / village square^1^ | Distribution of SMC at a specific location within village | Easier access than health facility or a single central point in the community for those who are far.  Usually high visibility and access to large crowds.  People are familiar with this approach for events like food distribution. | Large crowds can result in interruptions, disturbances, and conflict.  May be inaccessible to children or caretakers with disabilities.  Usually for distribution of food or goods, which may upset people if they feel tricked. |  |
| Nyumba kumi leaders | Provision of SMC to leaders of Nyumba Kumi where there are children hard to reach otherwise | Provision of SMC directly to households.  Easy to reach individuals living with disabilities and those that can’t go elsewhere or don’t trust people outside their Nyumba Kumi. | Training people that may have limited health / drug adminsitration experience.  Identifying Nyumba Kumi with children likely to be missed. |  |
| *Other strategies mentioned rarely* | | | | |
| At the play ground | Distribution of SMC at play ground | Reach children missed at schools or at home.  Large number of children in the same place. | Primary caretakers may not be present. |  |
| Train members of pastoralist kraals on distribution | Train focal points among pastoralists that migrate and provide with stock of SMC to distribute in their kraal | Pastoralist children receive SMC while on the move | Training focal points that may have limited health / drug administration experiences.  Limited monitoring of distribution. |  |
| Car parks (stages) | Administer SMC at car parks (stages) in town. | More likely to reach street children and children of people traveling a lot. | Crowding at the stages.  Some street children may not have their caretakers with them. |  |

**Appendix 2. Questions raised about SMC by community members**

- Why is SMC not administered to individuals outside the age group 3 – 59 months old (new-borns; older children; adults; the elderly)? Why are pregnant people not eligible?
- How do we know SMC is not poison?
- Can SMC be provided as an injection, a syrup, and a tablet?
- Can SMC be given if the child is ill?
- We used to take Fansidar for prevention and/or treatment of malaria, and then it went away. Why did that happen? And why is Fansidar being brought back now as a part of SMC?
- Is this going to lead to drug resistance later on?
- How long will SMC provide immunity to the child?
- What are the side effects of SMC?
- Can SMC be given on an empty stomach? If not, will SMC be accompanied by food?
- What happens if a child missing one of the SMC doses?
- What happens if a child takes a higher dosage than prescribed?
- How will you make sure there is enough drug for everybody?
- What happens after four months of SMC administration? What will protect us against malaria, especially where malaria is perceived as endemic?
- Who will administer SMC? *(some members did not trust CHPs to be able to correctly calculate the medication dose)*
- Does SMC work against all types of malaria?
- Sometimes drugs don’t work. Is there another approach like mosquito nets that you will provide?
